# Half of alcohol, drug, and self-harm presentations cannot be identified in coded emergency department data: a diagnostic accuracy study of a large language model

**DOI:** 10.64898/2026.08.26.26361443

**Authors:** Chris Humphries, Jonathan Brett, Franz Gruber, Edward James, Tuesday I McKendrick, Kimberley C McNairn, Anna Miell, Rachel O’Brien, Fahrurrozi Rahman, Lisa Schölin, Morven Stewart, Arlene Casey

## Abstract

**Objective:** To measure the accuracy of clinical coding, clinician review, and a locally deployed large language model (LLM) in identifying alcohol, drug, and self-harm involvement in emergency department (ED) attendances, and quantify prevalence.

**Design:** Two-phase diagnostic accuracy study. In a validation week, the identification strategies were assessed against a conflict-adjudicated reference standard (n=2,256); the LLM was then applied to n=105,096 annual attendances at the same site.

**Setting:** UK Type 1 Emergency Department treating patients ≥16yrs.

**Main outcome measures:** Prevalence quantification compared with the reference standard; sensitivity, specificity, and balanced accuracy of each strategy; monthly identification rates and adjusted annual prevalence.

**Results:** The reference standard identified 12.1% of attendances as involving alcohol, drugs, or self-harm (coding 6.0%; clinician 10.0%, LLM 15.6%). LLM balanced accuracy matched or outperformed clinician review in all three domains (alcohol 0.942 v 0.930, p=0.635; drug 0.959 v 0.791, p<0.001; self-harm 0.982 v 0.908, p=0.004). Coding recorded 1.07 domains per identified patient against 1.32 in the reference standard. Adjusted annual prevalence corresponded to 12,890 domain involvements per year not identifiable in coded data. Subdomain classification found at least 81.6% of self-harm attendances required medical assessment for injury or overdose before psychiatric review.

**Conclusions:** Clinical coding identified fewer than half of presentations involving alcohol, drugs, and self-harm and rarely captured co-occurring domains; under-recording was present across a full year. A locally deployed LLM generated more complete structured data from existing clinical text within NHS infrastructure, at a scale which is not feasible for manual review.

**What is already known on this topic:**

- Coded emergency department data underpin national surveillance, commissioning, research, and service planning, but the size and direction of their error for specific conditions have not been measured against a validated reference standard in an unselected population.

**What this study adds:**

- In a UK emergency department, clinical coding identified 6.0% of attendances as involving alcohol, drugs, or self-harm against an adjudicated reference-standard prevalence of 12.1%.
- In blinded head-to-head comparison, a locally deployed, open-source large language model achieved similar or higher balanced accuracy than clinician review. Applied to 105,096 attendances over 12 months, it identified 14.3% of attendances against 4.1% by coding, in every month of the year.

**How this study might affect research, practice, or policy:**

- Prevalence estimates and service planning based on coded emergency department data are likely to understate both the burden and the co-occurrence of these conditions. The majority (81.6%) of classified self-harm presentations required medical assessment for injury or overdose; only 18.4% did not, a figure directly relevant to mental health emergency-care streaming. Locally deployed open-source language models offer a scalable, governance-compatible method for identifying these presentations from existing free text.

## Introduction

Each year there are almost 30 million attendances at Emergency Departments (EDs) across the UK’s four health systems.[1-4] For each attendance, clinical codes are assigned by the treating clinician under time pressure and are not typically reviewed by a professional coder.[5] These codes underpin national surveillance, commissioning, epidemiological research, and, increasingly, the training data for clinical artificial intelligence.[6] Yet only around 6% of the information generated in a clinical consultation is entered as a structured (coded) field which can conceivably be used in datasets; the remainder is held as free text that is not routinely processed at scale.[7] Use of coded data therefore assumes that coded prevalence approximates true prevalence and that error is random and modest. Neither assumption has been tested against a validated reference standard in an unselected emergency population, nor examined over time.

Quantifying under-coding requires a domain in which ground truth can be independently established and in which under-ascertainment is both plausible and consequential. Presentations relating to alcohol, drugs, and self-harm meet these criteria. They are also a major public-health and policy priorities in their own right, which makes accurate emergency-care data on them especially consequential.[8-11] They are common, frequently co-occurring, and carry high morbidity and mortality.[12] They are also among the conditions most likely to be under-coded, because their involvement in a presentation is often secondary, implicit, or documented only in free-text notes. Involvement may also be under-recorded where a presentation is sensitive or stigmatised.[13] The need for better data to improve care in these domains is established.[12,14]

Data quality has direct impacts on policy. Poor-quality data lead to inaccurate estimates of activity and population need, and hence to misdirected resources. The UK government has committed £343 million to establish specialist mental health crisis centres, with pilots underway in ten trusts.[15-17] This investment assumes that a meaningful portion of mental health presentations can be identified at triage and streamed accordingly. The Royal College of Psychiatrists has warned that separating mental health from physical health emergency care risks missing the frequent co-occurrence of conditions.[17] The multi-domain prevalence data from unselected ED populations needed to resolve this debate do not exist.

Large language models (LLMs) can extract structured clinical information from free text with high accuracy,[18] and there is a clear case for applying them within health systems to generate structured data for systems intelligence, and to extract data from free text notes that cannot be shared with researchers directly due to privacy concerns.[12,14,18-20] Almost all existing evaluations have used curated datasets (for example, benchmark corpora or de-identified research extracts) rather than real, unedited clinical documentation.[21,22]

To assess whether these methods can unlock new data insights in emergency care, we conducted a two-phase diagnostic accuracy study in an unselected UK ED population. In a validation week including every attendance, we aimed to (1) quantify the gap between coded and true prevalence of alcohol, drug, and self-harm involvement against a conflict-adjudicated reference standard, and (2) measure the diagnostic accuracy of routine coding, blinded clinician review, and a blinded, locally deployed open-source LLM, including their capture of multi-domain co-occurrence and clinically relevant subdomains. We then applied the LLM to all 105,096 attendances over 12 months at the same site, to (3) determine whether the coding gap observed in the validation week persisted across a full year, and (4) estimate annual prevalence in each domain after correction for the LLM’s quantified error.

## Methods

### Study design and setting

Diagnostic accuracy study conducted in two phases at the Royal Infirmary of Edinburgh Emergency Department, a UK Type 1 department treating patients aged 16 and older, reported against STARD-AI,[23] with prevalence components additionally reported against STROBE[24] (Appendix 4). Phase 1 (validation week) established the conflict-adjudicated reference standard and assessed all three index strategies across every attendance (n=2,256) during a single index week in February 2024; no exclusion criteria were applied. The two-phase design is summarised in Supplementary Figure S1. Phase 2 (deployment year) applied the same LLM index test, with accuracy characterised in phase 1, to all 105,096 consecutive attendances between 1 January and 31 December 2024 at the same site; the validation week falls within this period.

### Ethical approval

This study originated as a locally registered clinical audit examining whether routine coding accurately reflected the true prevalence of alcohol, drug, and self-harm presentations, a question arising directly from service improvement work. The recognition that LLMs might offer a scalable solution to the under-ascertainment problem identified by the audit emerged subsequently, and the expanded study was developed accordingly. Transfer of the audit dataset for LLM evaluation within DataLoch (a partnership between the University of Edinburgh and NHS Lothian) was approved by the NHS Lothian Caldicott Guardian; subsequent method development and analysis of the data were conducted under existing NHS Lothian DataLoch information governance approvals.

### Classification strategies

Three strategies were compared for identifying the presence of alcohol, drug, or self-harm involvement in attendances. Each of alcohol, drug and self-harm is referred to as a domain, and an attendance could involve more than one (multi-domain). Domains were not mutually exclusive. Full definitions, including subdomain criteria are in Supplementary Appendix 1. Within each positive domain we also classified clinically relevant subdomains (for alcohol and drugs: intoxication, dependence, and related injury; for self-harm: intent and method). Homelessness was recorded as a patient-level social characteristic rather than a top-level domain.

#### Coding

Each attendance carried one structured triage code and a single diagnosis code, both assigned by the treating team in the ED at the point of care (using an Emergency Care Dataset-derived coding system)[5]. All triage and diagnostic codes with any plausible association with the three domains were identified. To distinguish attendances that coding missed entirely from those flagged under the wrong domain, all code-flagged cases additionally underwent clinician free-text review, forming a hybrid comparator (coding plus review) reported alongside the three primary strategies.

#### Clinician review

All 2,256 records were screened to identify attendances with any plausible association with alcohol, drugs, or self-harm (n=287 flagged), which subsequently underwent detailed data extraction using a structured audit tool. The classifications used in all reference-standard and accuracy analyses were made by a senior emergency medicine clinician (KCM; post-graduation year 8 [PGY8]), adopted after two PGY4 raters (TIM and MS) did not reach the pre-specified reliability threshold (inter-rater reliability in a 25% sample (threshold Krippendorff’s α ≥0.800)[25] reached α=0.795 despite additional training and repeat extraction, and review by a senior emergency medicine clinician (CH; PGY13) identified systematic classification errors). We therefore asked the PGY8 clinician to reclassify all 287 records, and used these classifications throughout.

#### LLM review

All 2,256 records were independently processed by Llama 3.1 70B Instruct (Meta; a 70-billion-parameter, instruction-tuned general-purpose language model, representative of the pre-agentic generation of LLMs that preceded the more autonomous, tool-using systems now commonly characterised as AI agents). The review used only the free-text triage and clinical note components, with no structured data fields. The LLM was deployed locally using vLLM (version 0.6.0) across two NVIDIA A100 GPUs within the secure data environment.[26] Inference used deterministic decoding (temperature 0), a maximum sequence length of 8,192 tokens, and a maximum generated output of 20 tokens per task. Three binary classification tasks were performed independently for each record (alcohol, drug, and self-harm involvement, defined as in Supplementary Appendix 1), each constrained to a single binary output, with the model instructed to assign a positive classification only where the record clearly supported it and zero when uncertain. The model was not fine-tuned, retrieval-augmented generation (providing the model with external reference text) was not used, and identical prompts and configuration were applied across the complete dataset (full prompt contract in Supplementary Appendix 2). The model was blinded to clinician outputs and clinical codes. Processing time was approximately 1.4 seconds per record.

### Reference standard

Where clinician and LLM agreed, this was accepted as the reference standard. Any disagreements were adjudicated by a PGY13 emergency medicine clinician subspecialised in clinical toxicology (CH), who reviewed the same text fields and made a final determination. The reference standard was therefore this adjudicated standard rather than PGY8 clinician review alone; clinician review is reported as one of the comparator strategies, which is why it can be outperformed.

### Outcomes

The phase 1 primary outcome was the prevalence of alcohol, drug, and self-harm involvement as determined by the reference standard, compared with the prevalence identified by clinical coding, clinician review, and LLM review. Phase 1 secondary outcomes were: (1) diagnostic accuracy of each strategy (balanced accuracy, sensitivity, specificity, PPV, NPV, F1); (2) multi-domain co-occurrence among reference-standard positive cases; (3) subdomain phenotyping performance (conditional accuracy for binary subdomains; unweighted Cohen’s kappa for multi-class self-harm classification, with linearly weighted kappa reported in Supplementary Table S1). Accuracy metrics for subdomains were assessed within the 287 clinician-subdomain-reviewed cases (Supplementary Appendix 1).

Phase 2 (comparison over 12 months of data) outcomes were: (1) monthly identification rates by coding and by the LLM; (2) annual adjusted prevalence per domain with confidence intervals; (3) absolute counts of attendances not identifiable from coded data; (4) consistency of the two corrections across the year.

### Statistical analysis

For phase 1, balanced accuracy was used as the principal accuracy metric because it weights sensitivity and specificity equally, is invariant to class prevalence, and is not inflated by the large true-negative majority. Paired comparisons of sensitivity and specificity between strategies were performed using McNemar’s test with Bonferroni correction for three co-primary comparisons (α=0.0167).[27] Differences in balanced accuracy were assessed using non-parametric bootstrap resampling (10,000 iterations, percentile method) for 95% confidence intervals.[28] For multi-class self-harm classification, a majority of confusion matrix cells contained zero counts; kappa values therefore likely understate the true magnitude of between-rater differences.[29] The significance threshold for secondary analyses was defined as p<0.05. Analysis was performed in R 4.5.0.

For phase 2, the identical LLM, prompts, and configuration processed one year of attendances, blinded to clinical codes throughout. Coded domain flags for the deployment year were derived by a deterministic mapping of each attendance’s triage and diagnosis code combination to the three domains; where a combination was ambiguous between domains, the mapping could assign more than one flag. The primary phase 2 analysis was the directly observed monthly gap between coded and LLM-identified prevalence. Apparent LLM prevalence was adjusted using the Rogan–Gladen method with the sensitivity and specificity characterised in phase 1, with bootstrap confidence intervals propagating test-characteristic uncertainty.[30] The same Rogan–Gladen adjustment was also applied to the coding data, using the sensitivity and specificity of coding estimated in phase 1. Stability of the coding gap was assessed descriptively as the monthly ratio of LLM-identified to coded prevalence. Applying these corrections across the deployment year assumes that the sensitivity and specificity measured during the validation week remained stable over time. The assumption is supported by the use of the same site, documentation system, model, prompts, and configuration throughout the study.

### Patient and public involvement

Following our local audit, PPI was conducted with the Scottish Association for Mental Health, the Scottish Drugs Forum, and VoxScotland. Participants emphasised that emergency care data need to capture ‘why’ patients attend, not just diagnostic codes, leading us to evaluate subdomain performance in order to describe the sub-types of each presentation (for example the nature of substance use, or the intent and method of self-harm). External method development or data processing by a commercial company was identified as unacceptable, necessitating locally deployed open-source models. Engagement took the form of structured discussions with these third-sector organisations, which represent people with lived experience of mental health, drug, and alcohol harm; participants advised on the value of free-text data and on acceptable models of data access. Sessions were facilitated by charity session-managers, and participants were compensated with vouchers.

## Results

### Validation week (phase 1)

#### Participants and reference standard

All 2,256 attendances in the validation week were classified by all three strategies, with no exclusions. The clinician and LLM classifications agreed in full for 1,999 attendances (88.6%); the remaining 257 (11.4%) disagreed in at least one domain and were adjudicated (Supplementary Table S2). The reference standard identified 274 attendances (12.1%) as involving alcohol (175; 7.8%), drugs (91; 4.0%), or self-harm (95; 4.2%); domains were not mutually exclusive.

#### Identification by clinical coding

Coding identified 136 attendances (6.0%) across the three domains (Table 1). On free-text review of these 136 cases, 114 (5.1% of all attendances) were confirmed as involving at least one domain and 22 were false positives. Review also reassigned domains within the flagged cases: drug flags fell from 57 to 31, while alcohol flags rose from 52 to 74 and self-harm from 37 to 67. Against the reference standard, coding identified 30% of alcohol involvement (coded 2.3% v 7.8%), 63% of drug involvement (2.5% v 4.0%), and 39% of self-harm involvement (1.6% v 4.2%) (Figure 1A).

**Table 1.** Prevalence of alcohol, drug, and self-harm involvement by identification strategy (n=2,256)

| Strategy | Alcohol n (%) | Drugs n (%) | Self-harm n (%) | Any domain n (%) |
| --- | --- | --- | --- | --- |
| **Coding (raw)** | 52 (2.3) | 57 (2.5) | 37 (1.6) | 136 (6.0) |
| **Coding + clinician free-text review** | 74 (3.3) | 31 (1.4) | 67 (3.0) | 114 (5.1) |
| **Clinician free-text review of all notes** | 159 (7.0) | 54 (2.4) | 87 (3.9) | 225 (10.0) |
| **LLM review of all notes** | 157 (7.0) | 243 (10.8) | 103 (4.6) | 353 (15.6) |
| **Reference standard** | 175 (7.8) | 91 (4.0) | 95 (4.2) | 274 (12.1) |

*Footnote: Coding + clinician free-text review = clinician free-text review of the 136 code-flagged attendances, removing false positives and reassigning domain flags where the code did not match the documented clinical picture. Domains not mutually exclusive. Abbreviations: LLM, Large Language Model.*

**Figure 1.**
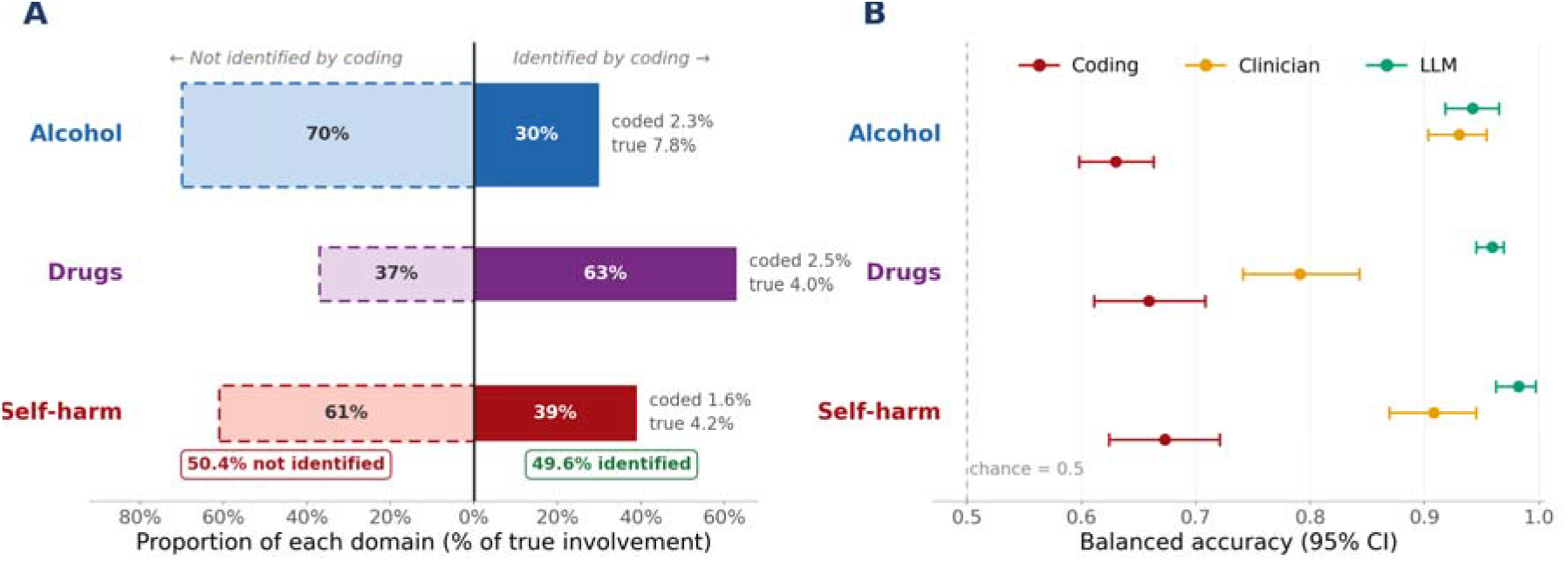
Identification of each domain by routine coding, and diagnostic accuracy of the three strategies (validation week). A) Diverging bars: proportion of each domain not identified (left) versus identified (right) by routine coding; bar widths proportional to reference-standard prevalence. B) Balanced accuracy with 95% bootstrap confidence intervals (10,000 resamples), 0.5 representing chance. Full per-metric performance profiles are in Supplementary Table S3.

#### Diagnostic accuracy

The LLM matched or exceeded the balanced accuracy of clinician review in all three domains (Table 2, Figure 1B). On McNemar testing (Bonferroni-corrected α=0.0167), LLM sensitivity was higher than coding in all three domains (all p<0.001) and higher than the clinician for drugs (p<0.001) and self-harm (p=0.004), with no difference for alcohol (p=0.635); clinician specificity was higher than the LLM for drugs (p<0.001). Coding sensitivity ranged from 0.263 to 0.347, with specificity of 0.988 or above (Supplementary Tables S3 and S4).

**Table 2.** Diagnostic accuracy by strategy and domain (n=2,256)

|  | Alcohol Coding | Alcohol Clinician | Alcohol LLM | Drug Coding | Drug Clinician | Drug LLM | Self-harm Coding | Self-harm Clinician | Self-harm LLM |
| --- | --- | --- | --- | --- | --- | --- | --- | --- | --- |
| **Bal. accuracy** | .630 | .930 | .942 | .659 | .791 | .959 | .673 | .908 | .982 |
| **95% CI** | .598–.663 | .903–.954 | .918–.965 | .611–.708 | .741–.843 | .945–.969 | .624–.721 | .869–.945 | .962–.997 |
| **Sensitivity** | .263 | .863 | .886 | .330 | .582 | .989 | .347 | .821 | .968 |
| **Specificity** | .997 | .996 | .999 | .988 | .999 | .929 | .998 | .996 | .995 |
| **PPV** | .885 | .950 | .987 | .526 | .981 | .370 | .892 | .897 | .893 |
| **NPV** | .941 | .989 | .990 | .972 | .983 | .999 | .972 | .992 | .999 |

Footnote: 95% CIs for balanced accuracy from non-parametric bootstrap (10,000 resamples). F1 scores, full confusion matrices, and McNemar's test results in Supplementary Tables S3 and S4.

#### Characterisation and correction of LLM error

The LLM’s principal error was in the drug domain, where sensitivity of 0.989 was accompanied by positive predictive value of 0.370. Post hoc review identified three sources of false positives: adverse medication reactions, composite triage set text populating text fields without confirming genuine drug involvement (e.g. “drug/alcohol intoxication/withdrawal”), and therapeutic benzodiazepine administration for alcohol withdrawal. A refined prompt tested post hoc reduced false positives but increased false negatives from 1 to 42, worsening balanced accuracy from 0.959 to 0.755 (Supplementary Table S5); the primary prompt with statistical adjustment was therefore retained for phase 2, with Rogan-Gladen adjustment (a correction method using a test’s measured sensitivity and specificity to convert an observed prevalence into an estimate of the true prevalence) using quantified performance from phase 1.

#### Multi-domain co-occurrence

Among the 274 reference-standard positive patients, 73 (26.6%) had involvement of more than one domain (Supplementary Table S6; Supplementary Figures S2 and S3). The most common pattern was single-domain alcohol involvement (123; 44.9%), followed by isolated self-harm (41; 15.0%) and isolated drugs (37; 13.5%); all three domains co-occurred in 14 patients (5.1%). Of the 95 patients with self-harm, 54 (56.8%) had co-occurring alcohol or drug involvement. Coding could be deterministically mapped to generate a mean of 1.07 domain flags per identified patient, compared with 1.32 for the reference standard and 1.42 for the LLM (Supplementary Table S7).

#### Subdomain classification

Within the 287 clinician-reviewed cases, the LLM classified subdomains with balanced accuracy similar to or higher than the clinician reviewer (Figure 2A; Supplementary Table S8), including higher agreement with the reference standard for self-harm intent (κ=0.656 [0.514 to 0.787] v 0.442 [0.325 to 0.559]; p=0.013). Both strategies performed poorly on drug subdomains, in opposite directions (Supplementary Tables S4 and S8). Subdomain classification was available for 87 of 95 reference-standard self-harm cases; the remaining 8 were identified by the LLM only and were not subject to full blinding subdomain classification by a clinician. Of the 87 classified cases, 16 (18.4%) had no injury or overdose requiring medical assessment before psychiatric review (i.e. suicidal ideation only or undisclosed intent staff concern only). Fourteen of these patients had alcohol or drug co-involvement in their presentation (Figure 2B).

**Figure 2.**
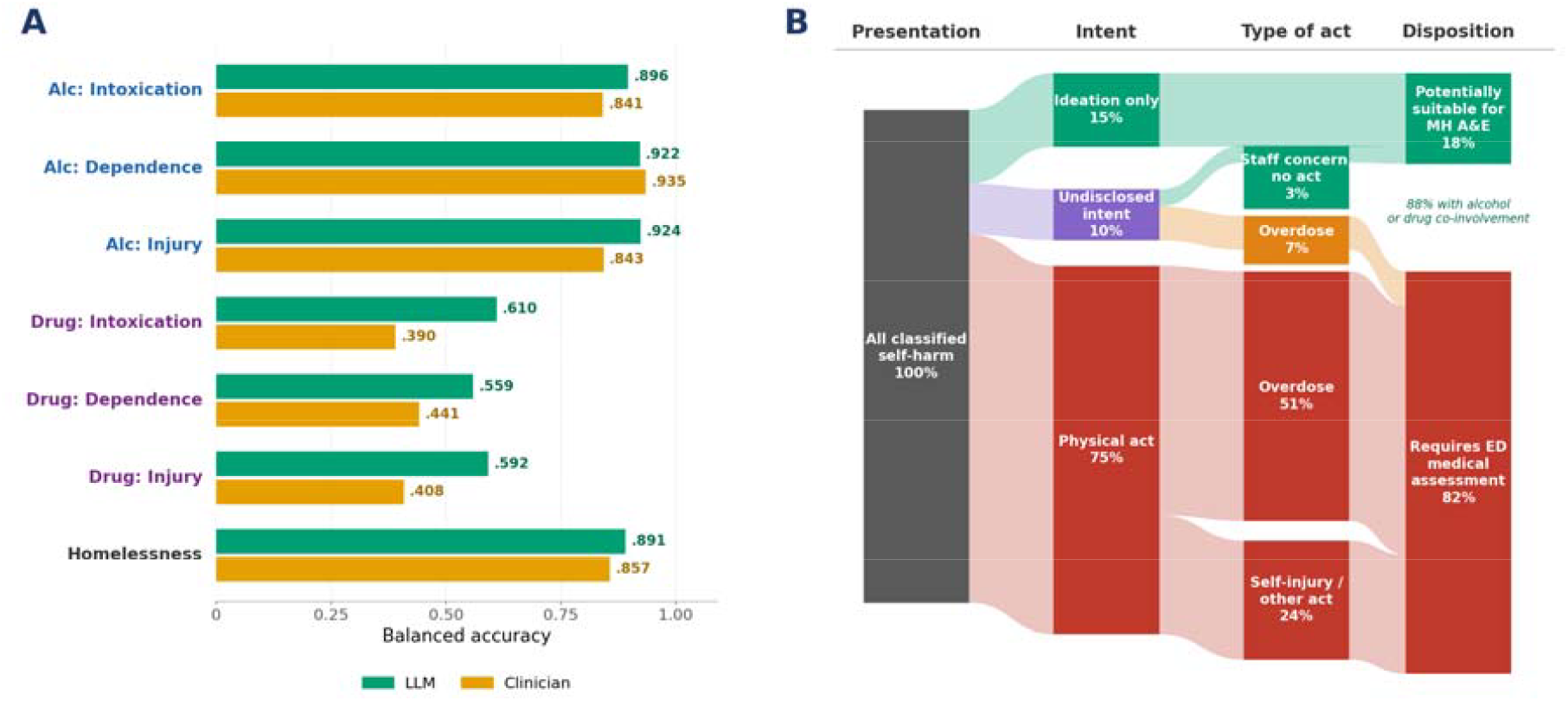
Subdomain classification performance and the self-harm care pathway (validation week). A) Subdomain balanced accuracy, clinician versus LLM (exploratory; no formal pairwise tests; full metrics in Supplementary Table S8). B) Care pathway of the 87 classified self-harm presentations from intent through type of act to disposition. The ideation/no-act group is labelled ‘potentially suitable’ because 14 of the 16 had alcohol or drug co-involvement, which may preclude direct streaming to a mental health facility. Ribbon widths and final nodes are proportional to patient numbers; some node heights are enlarged to fit their labels.

### Deployment year (phase 2)

#### Monthly identification rates

The LLM classified all 105,096 attendances between 1 January and 31 December 2024 (approximately 1.4 seconds per record; approximately 42 hours of inference in total), blinded to clinical codes. Coding identified 4,352 attendances (4.1%) as involving at least one domain and the LLM identified 15,032 (14.3%); 205 attendances were identified by coding but not the LLM. The difference was present in every month and every domain (Figure 3, Supplementary Table S9). Monthly identification by coding ranged from 3.4% to 4.9% of attendances (any domain), attendances identified by the LLM but not by coding added a further 9.8 to 11.3 percentage points (any domain), and the LLM identified roughly three to four times as many attendances as coding each month (ratio 3.0 to 3.9) throughout. The coded proportion was lower throughout the year than in the validation week (6.0%), while the LLM proportion was similar (15.6% in the validation week).

**Figure 3.**
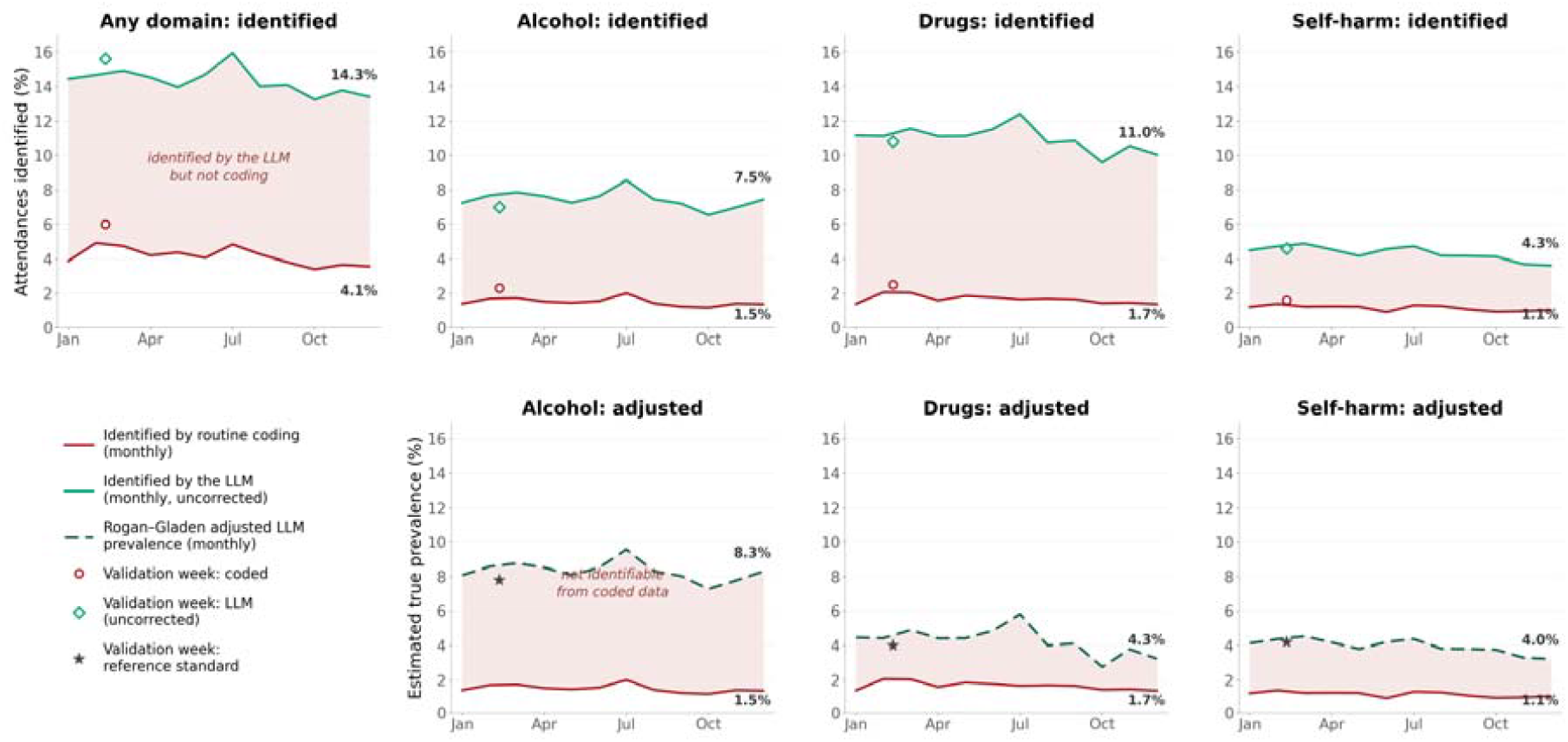
Monthly identification and adjusted prevalence across the deployment year (n=105,096). Upper panels: proportion of attendances identified as involving each domain by routine coding and by the LLM; shaded area identifies the gap. Lower panels: monthly prevalence after Rogan–Gladen adjustment of the LLM series; the shaded area represents attendances whose domain involvement is not identifiable from coded data. Annual adjusted values with confidence intervals are in Table 3; the any-domain composite cannot be Rogan–Gladen adjusted (see the Table 3 footnote). The markers represent a single week while the lines are calendar-month proportions. Direct labels show annual values.

#### Adjusted annual prevalence

After Rogan–Gladen adjustment using phase 1 test characteristics, annual prevalence was 8.3% (95% CI 7.8 to 8.9) for alcohol, 4.3% (3.1 to 5.4) for drugs, and 4.0% (3.6 to 4.3) for self-harm, against coded rates of 1.5%, 1.7%, and 1.1% respectively (Table 3). These estimates correspond to 7,181 alcohol-involved, 2,746 drug-involved, and 2,963 self-harm-involved attendances per year, 12,890 domain involvements in total, that could not be identified from this department’s coded record. The adjusted estimates were consistent with the validation-week reference values in all three domains (7.8%, 4.0%, and 4.2%); for drugs, the apparent prevalence of 11.0% adjusted to 4.3%. Monthly adjusted rates, with the corresponding coded rates, are shown in the lower panels of Figure 3.

**Table 3.** Annual identification and adjusted prevalence by domain across the deployment year (n=105,096)

| Domain | Coded n (%) | LLM n (%) | LLM adjusted % (95% CI) | Estimated true n | Not identifiable from codes, n |
| --- | --- | --- | --- | --- | --- |
| Alcohol | 1,565 (1.5) | 7,839 (7.5) | 8.3 (7.8–8.9) | 8,746 | 7,181 |
| Drugs | 1,743 (1.7) | 11,550 (11.0) | 4.3 (3.1–5.4) | 4,489 | 2,746 |
| Self-harm | 1,202 (1.1) | 4,547 (4.3) | 4.0 (3.6–4.3) | 4,165 | 2,963 |
| Any domain | 4,352 (4.1) | 15,032 (14.3) | – | – | – |

*Adjusted values by the Rogan–Gladen method using phase 1 sensitivity and specificity; 95% CIs from 10,000 bootstrap resamples of the phase 1 cohort with binomial sampling of the year counts. Estimated true n = adjusted prevalence × 105,096. Domains not mutually exclusive; any-domain row shows observed counts. No adjusted estimate is shown for the any-domain composite because Rogan–Gladen correction requires a single sensitivity and specificity, which are not defined for a union of three domains with differing test characteristics. CIs computed against the phase 1 patient-level data.*

## Discussion

### Principal findings

Assessed against a conflict-adjudicated reference standard covering every attendance in one week, clinical coding identified fewer than half of ED presentations involving alcohol, drugs, or self-harm (6.0% v 12.1%), frequently assigned the flagged cases to the wrong domain, and rarely recorded co-occurring domains. A locally deployed open-source LLM identified these presentations with balanced accuracy similar to or higher than blinded clinician review. Applied to 105,096 attendances over a full year, it identified 14.3% of attendances against 4.1% by coding, with the discrepancy apparent across the year. England’s Emergency Care Data Set permits secondary diagnoses, but our experience suggests most attendances receive only a primary diagnosis even when several problems are present, so this information is lost by design at some sites and by practice at others. The LLM approach recovers it from what clinicians already write, without requiring any change in clinical behaviour.

### Implications for mental health crisis centres

Specialist mental health crisis centres, backed by £343 million of investment, assume that mental health presentations can be identified and streamed at triage.[15,17] Our data qualify this in three ways: coding identified 1.6% of attendances as self-harm against a reference-standard 4.2%, so code-based triage would miss 61% (58/95) of the self-harm population; only 16 of 87 classified self-harm presentations (18.4%) had no injury or overdose requiring medical assessment (and 14 of those 16 had co-occurring substance involvement, leaving just 2.3% with no evidence of a potential physical care need nor substance co-involvement). Services designed for isolated mental health presentations would be mismatched to this case-mix.[17,31] Applied to the UK’s almost 30 million annual ED attendances (29.7 million in 2023-24),[1-4] a shortfall of this magnitude would leave several million domain involvements unidentified each year, though case-mix and coding differ across settings.

An analogous limitation applies to substance-use care: services that depend on accurately identifying alcohol- and drug-related attendances, such as alcohol care teams and emergency-department addiction liaison, would face the same shortfall, as coding identified only 2.3% of attendances as alcohol-related and 2.5% as drug-related against reference-standard rates of 7.8% and 4.0% respectively.

### Implications for research and surveillance

Research cohorts selected by coded domain will be incomplete and unrepresentative, and coded co-occurrence is too sparse to define cohorts by it (coding identified only 158 multi-domain attendances across the 105,096, against roughly 7,700 identified by the LLM). The same under-ascertainment plausibly affects other conditions documented mainly in free text. Subdomain extraction further widens what routine data can answer: classifying self-harm intent, distinguishing dependence from intoxication, or identifying homelessness at population scale has previously required bespoke prospective collection, and can now be measured retrospectively from records that already exist.

### From unstructured text to structured data within existing governance

Privacy concerns have generally prevented free-text notes from being provided to researchers within Trusted Research Environments. This study demonstrates an alternative: LLM extraction as a back-end process within existing governance, converting notes into structured datasets accessible through standard pathways.[19] The de-identified domain and subdomain flags carry no greater privacy risk than existing coded data, with higher sensitivity and more complete capture. Public survey and deliberative evidence indicate conditional support for using NHS data in this way within trusted research environments under the Five Safes framework.[32,33]

### Comparison with existing literature

To our knowledge this is the first evaluation of LLM extraction on real, unedited ED documentation in an operational NHS setting, and the first to measure coding performance continuously over a year against an extraction method of known test characteristics. Prior studies have typically used MIMIC-IV (a large, de-identified US critical-care research database)[34] or curated text.[35] For prevalence estimation, statistical adjustment of a high-sensitivity classifier[30] proved preferable to prompt optimisation that sacrifices sensitivity; retrieval-augmented generation[36,37] and emerging open-source pipelines[38,39] may improve precision and support multi-site replication.

### Limitations

This is a single-site study; exact figures will vary by site, case-mix, and documentation practice. The mechanism, time-pressured coding of complex presentations documented mainly in free text, is common to UK EDs, and our prior work in another setting found comparable discrepancies.[14] The full deployment year makes an atypical validation week an unlikely explanation, and a prospective multi-site validation is underway.

The reference standard is not fully independent of the tests under evaluation. Where the clinician and LLM agreed, that agreement was accepted without further review; errors shared by both would be incorporated into the reference standard, inflating the apparent accuracy of both. Coding did not contribute to the reference standard and is unaffected by this mechanism. A domain-specific caveat applies to the drug results: the reference standard was adjudicated by a clinician subspecialised in clinical toxicology. Part of the apparent shortfall in clinician review for the drug domain may therefore reflect ascertainment bias.

The deployment-year estimates assume that the test characteristics measured in the validation week held across the year. Site, documentation system, model, prompts, and configuration were constant; nevertheless, no within-year reference standard exists. The divergence between the coding-corrected and LLM-corrected annual estimates indicates that at least one method departed from its validation-week characteristics. This may reflect gradual shifts in coding behaviour, seasonal change relative to a February validation week, or differences between the year’s deterministic code-to-domain mapping and the week’s manual code list; these cannot be distinguished here.

Only one clinician performed the blinded top-level review and subdomain classification, so interrater reliability was not assessed. A drug-domain PPV of 0.370 would be unacceptable for clinical decision-making about an individual patient. For estimating prevalence, however, the LLM’s high sensitivity and predictable false positives allowed its error to be corrected, so the adjusted prevalence matched the reference standard (4.1% v 4.0%). Coding’s low sensitivity, by contrast, cannot be detected or corrected from routine data alone.

Ethical approvals for this study did not extend to linkage with demographic or socioeconomic datasets, or identification rates by age, sex, and deprivation. Whether coding quality and LLM accuracy vary by deprivation, ethnicity, age, or sex is an important equity question; subgroup analysis should form a component of future work.

### Conclusions

Clinical coding identified fewer than half of emergency department presentations involving alcohol, drugs, or self-harm and rarely recorded co-occurring domains; across 105,096 attendances the shortfall was present in every month of the year. A locally deployed open-source LLM matched blinded clinician review and generated more complete structured data from existing clinical text, within current NHS infrastructure and governance. Methods now exist to measure, and substantially reduce, this gap in routinely collected emergency care data.

## Supporting information

STARD-AI

STROBE

## Data availability

Patient-level data cannot be shared; the DataLoch extract used for this development project cannot be requested in its raw form. The derived structured data may be permitted to form part of future Trusted Research Environment projects, subject to ethical approvals.

## Author contributions

Conceptualization: CH, JB, AC; Data curation: CH, FG, AC; Formal analysis: CH, AC; Investigation: CH, JB, FG, TIM, KCM, AM, ROB, MS, AC; Methodology: CH, JB, AC; Project administration: CH; Resources: CH, EJ, AC; Software: CH, FG, AC, FR; Supervision: CH; Validation: CH, FG, AC; Visualization: CH; Writing – original draft: CH; Writing – review and editing: CH, JB, FG, EJ, TIM, KCM, AM, ROB, LS, MS, AC, FR

