## Supplemental for "Half of alcohol, drug, and self-harm presentations cannot be identified in coded emergency department data: a diagnostic accuracy study of a large language model"

#### **Supplementary Figure S1: Study design**

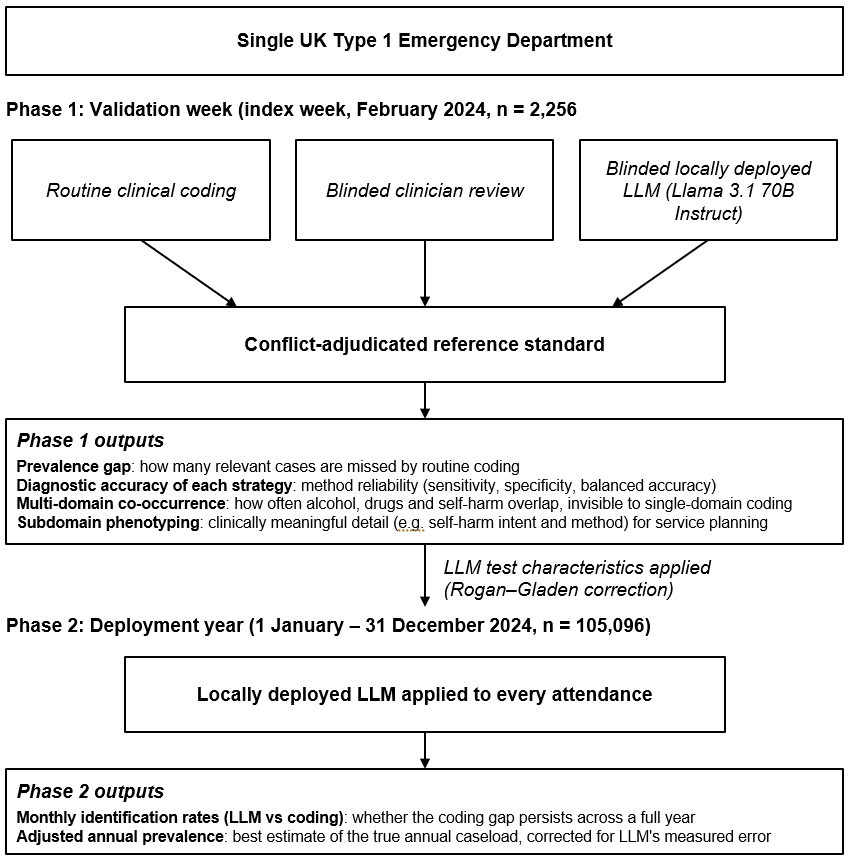

*Two-phase diagnostic accuracy study design. Phase 1 (validation week, February 2024; n=2,256 consecutive attendances) compared routine clinical coding, blinded clinician review, and a blinded, locally deployed large language model against a conflict-adjudicated reference standard, yielding the prevalence gap, diagnostic accuracy of each strategy, multi-domain co-occurrence, and subdomain phenotyping. Phase 2 (deployment year, 1 January to 31 December 2024; n=105,096 attendances) applied the LLM to every attendance to test whether the coding gap persisted across a full year and to estimate adjusted annual prevalence (Rogan-Gladen correction using the phase 1 test characteristics).*

#### **Supplementary Figure S2: Domain co-occurrence among reference-standard positive patients**

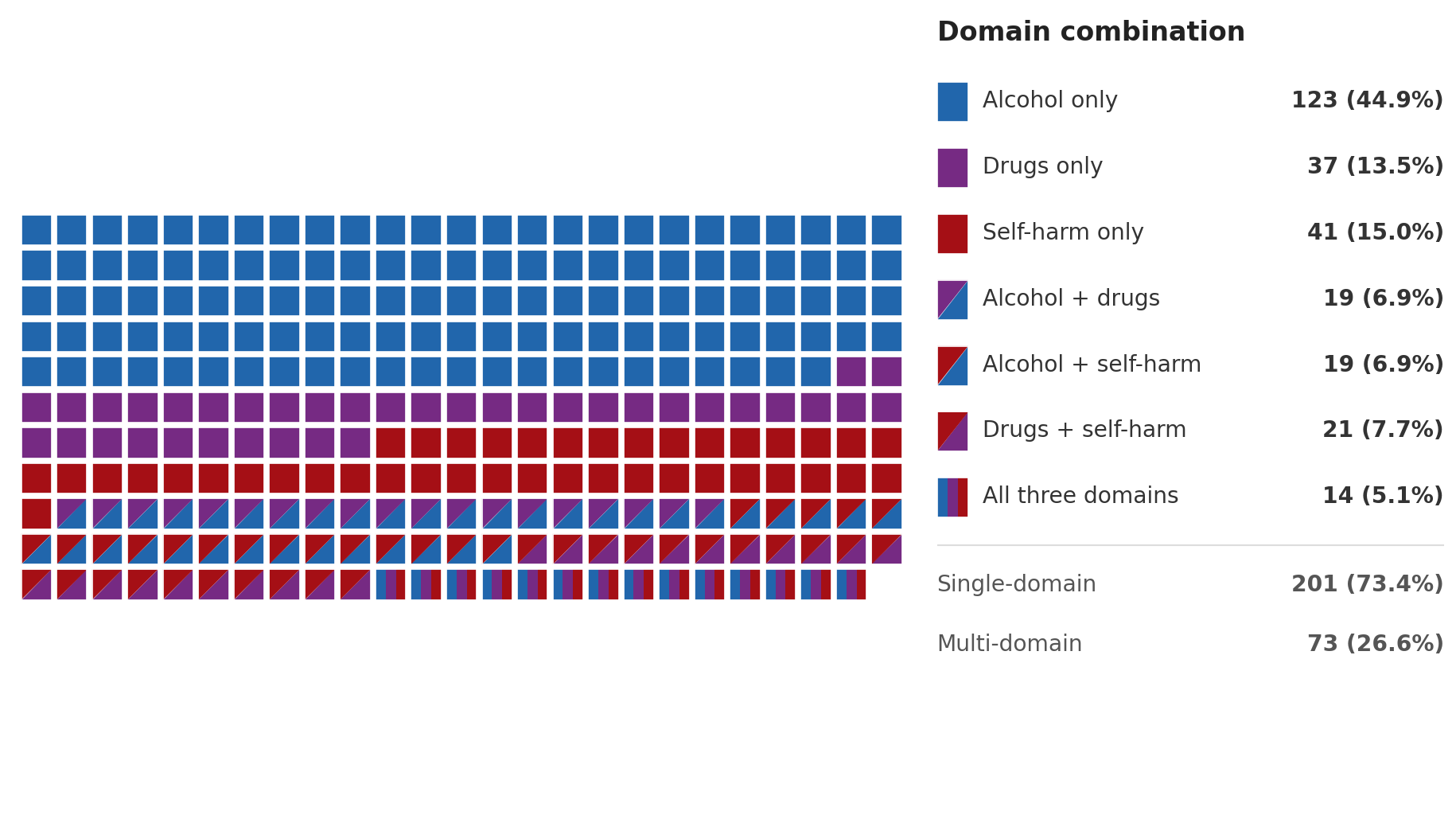

*Waffle plot of domain co-occurrence among the 274 affected patients; colour indicates domain combination (single-domain 201, 73.4%; multi-domain 73, 26.6%). Each square represents one patient; squares split diagonally (two domains) or striped (all three) indicate co-occurring domains.*

#### **Supplementary Figure S3: Domain co-occurrence (UpSet plot)**

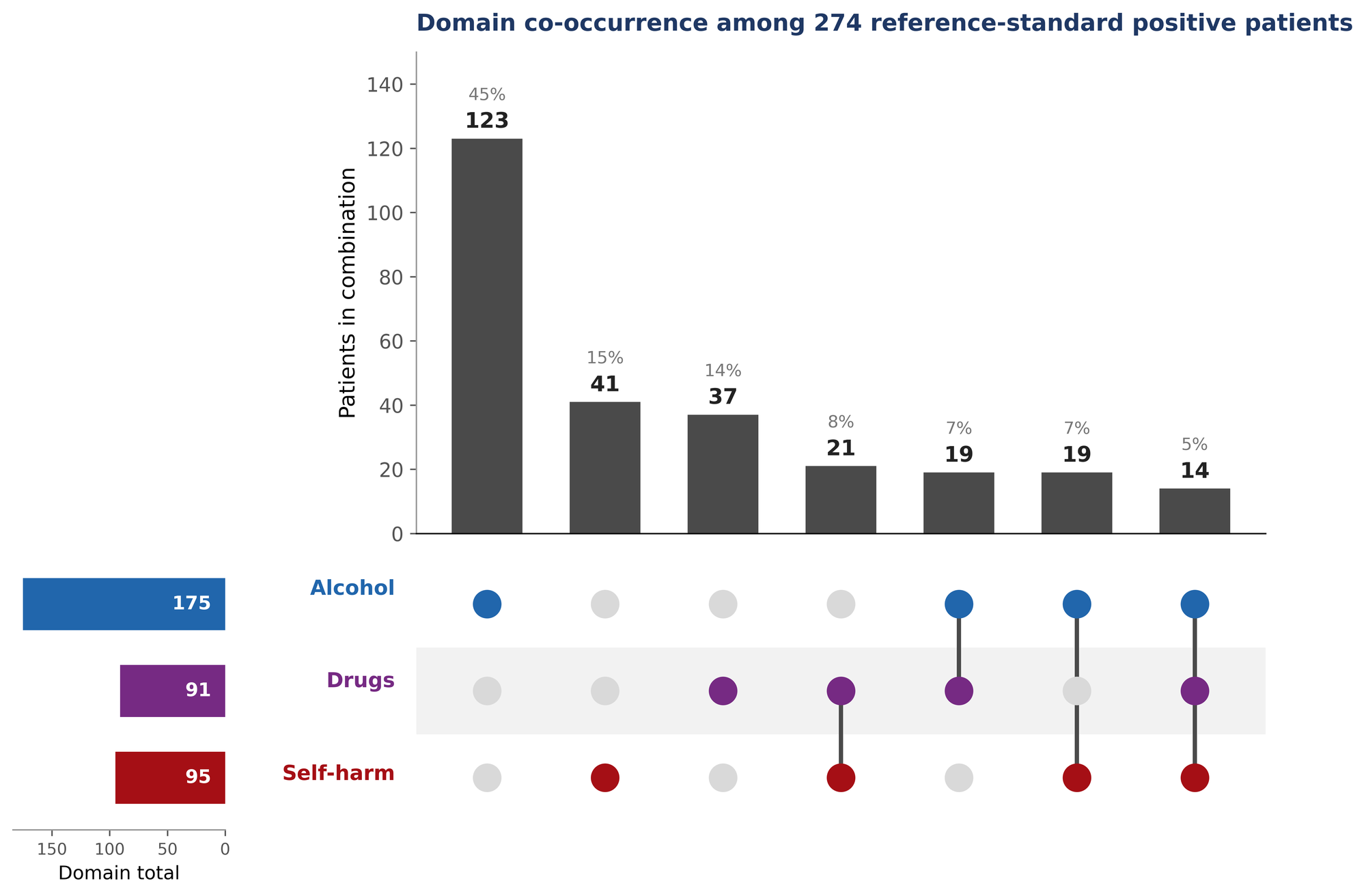

*UpSet plot of domain co-occurrence among the 274 reference-standard positive patients. Vertical bars show the number of patients in each domain combination (with the percentage of the 274); the dot matrix indicates which domains are involved; horizontal bars (lower left) show the total number of patients positive for each domain. The same data are shown as a waffle plot in Supplementary Figure S2.*

#### **Supplementary Table S1: Self-harm multi-class kappa**

| **Domain** | **Weighting** | **LLM κ [95% CI]** | **Clin κ [95% CI]** | **Diff [95% CI]** | **p** |
| --- | --- | --- | --- | --- | --- |
| Intent | Unweighted | 0.656 [0.514, 0.787] | 0.442 [0.325, 0.559] | 0.214 [0.027, 0.398] | .013 |
| Intent | Linearly weighted | 0.595 [0.423, 0.750] | 0.309 [0.155, 0.469] | 0.286 [0.043, 0.517] | .011 |
| Type | Unweighted | 0.785 [0.652, 0.898] | 0.690 [0.565, 0.809] | 0.094 [−0.086, 0.272] | .154 |
| Type | Linearly weighted | 0.699 [0.524, 0.860] | 0.600 [0.418, 0.770] | 0.100 [−0.153, 0.358] | .229 |

*Bootstrap CIs (10,000 resamples). Majority of confusion matrix cells contained zero counts; kappa may understate true differences. Uncorrected p-values.*

#### **Supplementary Table S2: Clinician vs LLM disagreements at adjudication (validation week)**

Of the 2,256 validation-week attendances, the clinician and LLM agreed on all three domains for 1,999 (88.6%); the remaining 257 (11.4%) disagreed on at least one domain and were adjudicated. Because an attendance can disagree in more than one domain, these 257 attendances contain 281 domain-level disagreements.

| **Domain** | **Both flagged** | **LLM only** | **Clinician only** | **LLM-only confirmed** | **Clinician-only confirmed** |
| --- | --- | --- | --- | --- | --- |
| **Alcohol** | 133 | 24 | 26 | 22 / 24 | 18 / 26 |
| **Drugs** | 53 | 190 | 1 | 37 / 190 | 0 / 1 |
| **Self-harm** | 75 | 28 | 12 | 17 / 28 | 3 / 12 |
| **Total** | - | 242 | 39 | 76 / 242 | 21 / 39 |

*‘Both flagged’ = domain flagged by both clinician and LLM. ‘LLM only’ / ‘Clinician only’ = flagged by one strategy but not the other. ‘Confirmed’ = adjudicated positive by the reference standard. Across all domains the LLM identified 76 adjudicated-true cases the clinician missed, versus 21 the other way. The drug domain is the exception: of 190 LLM-only flags, 153 were false positives (the low-precision behaviour, PPV 0.37, corrected by Rogan-Gladen adjustment).*

#### **Supplementary Table S3: Full confusion matrices, validation week (N=2,256)**

| **Domain** | **Strategy** | **TP** | **FP** | **FN** | **TN** | **Sens** | **Spec** | **PPV** | **NPV** | **F1** | **BA** |
| --- | --- | --- | --- | --- | --- | --- | --- | --- | --- | --- | --- |
| Alcohol | Coding | 46 | 6 | 129 | 2075 | .263 | .997 | .885 | .941 | .405 | .630 |
| Alcohol | Clinician | 151 | 8 | 24 | 2073 | .863 | .996 | .950 | .989 | .904 | .930 |
| Alcohol | LLM | 155 | 2 | 20 | 2079 | .886 | .999 | .987 | .990 | .934 | .942 |
| Drugs | Coding | 30 | 27 | 61 | 2138 | .330 | .988 | .526 | .972 | .405 | .659 |
| Drugs | Clinician | 53 | 1 | 38 | 2164 | .582 | .999 | .981 | .983 | .731 | .791 |
| Drugs | LLM | 90 | 153 | 1 | 2012 | .989 | .929 | .370 | .999 | .539 | .959 |
| Self-harm | Coding | 33 | 4 | 62 | 2157 | .347 | .998 | .892 | .972 | .500 | .673 |
| Self-harm | Clinician | 78 | 9 | 17 | 2152 | .821 | .996 | .897 | .992 | .857 | .908 |
| Self-harm | LLM | 92 | 11 | 3 | 2150 | .968 | .995 | .893 | .999 | .929 | .982 |

#### **Supplementary Table S4: McNemar’s test results (Bonferroni α=0.0167)**

| **Domain** | **Comparison** | **Test** | **A>B** | **B>A** | **χ²** | **p** | **Result** |
| --- | --- | --- | --- | --- | --- | --- | --- |
| Alcohol | LLM vs Clin | Sens | 22 | 18 | 0.23 | .635 | NS |
| Alcohol | LLM vs Code | Sens | 109 | 0 | 107.0 | <.001 | LLM superior |
| Drugs | LLM vs Clin | Sens | 37 | 0 | 35.0 | <.001 | LLM superior |
| Drugs | LLM vs Clin | Spec | 1 | 153 | 148.1 | <.001 | Clin superior |
| Drugs | LLM vs Code | Sens | 60 | 0 | 58.0 | <.001 | LLM superior |
| Self-harm | LLM vs Clin | Sens | 17 | 3 | 8.45 | .004 | LLM superior |
| Self-harm | LLM vs Code | Sens | 60 | 1 | 55.1 | <.001 | LLM superior |

*A>B = first strategy correct, second incorrect (discordant pairs). Only comparisons with significant or near-significant results shown; full table available from authors.*

#### **Supplementary Table S5: Drug domain prompt comparison**

| **Prompt** | **TP** | **FP** | **FN** | **TN** | **Sens** | **Spec** | **PPV** | **F1** | **BA** |
| --- | --- | --- | --- | --- | --- | --- | --- | --- | --- |
| Primary | 90 | 153 | 1 | 2012 | .989 | .929 | .370 | .539 | .959 |
| Refined | 49 | 62 | 42 | 2103 | .538 | .971 | .441 | .485 | .755 |

*The refined prompt reduced FP from 153 to 62 but increased FN from 1 to 42; every composite metric worsened. For prevalence estimation, Rogan–Gladen adjustment of the primary prompt is preferable.*

#### **Supplementary Table S6: Domain co-occurrence among reference-standard positive patients (n=274)**

| **Combination** | **Domains** | **n** | **% of affected** |
| --- | --- | --- | --- |
| Alcohol only | 1 | 123 | 44.9 |
| Self-harm only | 1 | 41 | 15.0 |
| Drugs only | 1 | 37 | 13.5 |
| Drugs + Self-harm | 2 | 21 | 7.7 |
| Alcohol + Drugs | 2 | 19 | 6.9 |
| Alcohol + Self-harm | 2 | 19 | 6.9 |
| All three | 3 | 14 | 5.1 |

*Domains not mutually exclusive. Combination counts sum to 274. See Supplementary Figure S2 for the waffle plot visualisation.*

#### **Supplementary Table S7: Information density, validation week**

| **Strategy** | **Patients flagged** | **Total domain flags** | **Flags per patient** |
| --- | --- | --- | --- |
| Clinical coding | 136 | 146 | 1.07 |
| Clinician review | 225 | 300 | 1.33 |
| LLM extraction | 353 | 503 | 1.42 |
| Reference standard | 274 | 361 | 1.32 |

#### **Supplementary Table S8: Subdomain performance (n=287)**

| **Subdomain** | **Clin Sens** | **Clin Spec** | **Clin BA** | **Clin F1** | **LLM Sens** | **LLM Spec** | **LLM BA** | **LLM F1** |
| --- | --- | --- | --- | --- | --- | --- | --- | --- |
| Alc: Intoxication | .955 | .727 | .841 | .926 | .928 | .864 | .896 | .936 |
| Alc: Dependence | .892 | .978 | .935 | .928 | .877 | .967 | .922 | .912 |
| Alc: Injury | .759 | .928 | .843 | .807 | .879 | .969 | .924 | .911 |
| Drug: Intoxication | .053 | .727 | .390 | .087 | .947 | .273 | .610 | .800 |
| Drug: Dependence | .000 | .882 | .441 | .000 | 1.00 | .118 | .559 | .571 |
| Drug: Injury | .200 | .615 | .408 | .235 | .800 | .385 | .592 | .615 |
| Homelessness | .733 | .982 | .857 | .733 | .800 | .982 | .891 | .774 |

*Conditional accuracy: assessed only among patients flagged by at least one strategy for the parent domain, representing a credible production pipeline.*

#### **Supplementary Table S9: Monthly prevalence by strategy, deployment year**

| **Month** | **Episodes** | **Any: coded % / LLM % (ratio)** | **Alcohol: coded % / LLM % (ratio)** | **Drugs: coded % / LLM % (ratio)** | **Self-harm: coded % / LLM % (ratio)** |
| --- | --- | --- | --- | --- | --- |
| Jan | 8,621 | 3.9 / 14.4 (3.7) | 1.4 / 7.3 (5.2) | 1.4 / 11.2 (8.1) | 1.2 / 4.5 (3.7) |
| Feb | 8,389 | 4.9 / 14.7 (3.0) | 1.7 / 7.7 (4.5) | 2.1 / 11.1 (5.4) | 1.4 / 4.7 (3.4) |
| Mar | 8,919 | 4.7 / 14.9 (3.2) | 1.7 / 7.9 (4.6) | 2.0 / 11.6 (5.7) | 1.2 / 4.9 (4.0) |
| Apr | 8,476 | 4.2 / 14.5 (3.5) | 1.5 / 7.6 (5.1) | 1.6 / 11.1 (7.0) | 1.2 / 4.5 (3.7) |
| May | 9,342 | 4.4 / 14.0 (3.2) | 1.4 / 7.3 (5.0) | 1.9 / 11.1 (6.0) | 1.2 / 4.2 (3.4) |
| Jun | 8,780 | 4.1 / 14.7 (3.6) | 1.5 / 7.6 (5.0) | 1.8 / 11.5 (6.5) | 0.9 / 4.6 (5.0) |
| Jul | 8,988 | 4.8 / 15.9 (3.3) | 2.0 / 8.6 (4.3) | 1.6 / 12.4 (7.5) | 1.3 / 4.7 (3.6) |
| Aug | 8,980 | 4.3 / 14.0 (3.3) | 1.4 / 7.5 (5.3) | 1.7 / 10.7 (6.4) | 1.3 / 4.2 (3.3) |
| Sep | 8,744 | 3.8 / 14.1 (3.7) | 1.2 / 7.2 (5.9) | 1.6 / 10.9 (6.6) | 1.1 / 4.2 (3.9) |
| Oct | 9,100 | 3.4 / 13.2 (3.9) | 1.2 / 6.5 (5.6) | 1.4 / 9.6 (6.8) | 0.9 / 4.1 (4.4) |
| Nov | 8,308 | 3.7 / 13.8 (3.8) | 1.4 / 7.0 (5.0) | 1.4 / 10.5 (7.3) | 1.0 / 3.7 (3.8) |
| Dec | 8,449 | 3.6 / 13.4 (3.8) | 1.4 / 7.4 (5.5) | 1.4 / 10.0 (7.4) | 1.0 / 3.6 (3.6) |

*Coded % = attendances identified by routine coding; LLM % = attendances identified by the LLM; ratio (in parentheses) = LLM % / coded %. LLM figures are LLM-identified proportions, consistent with Table 3 and Figure 3. Across the year, 205 attendances were identified by coding but not the LLM.*

### **Supplementary Appendix 1**

**Data Abstraction Definitions**

This appendix defines the data abstraction criteria used by clinician reviewers.

*Note: All reviewers were instructed to focus on correspondence and clinical notes from the index attendance dates only.*

| **Criterion** | **Sub-criterion** | **Definition** | **Options** |
| --- | --- | --- | --- |
| **Presentation of interest?** |  | Presentations of interest are any related to (i.e. current effects or consequences of) alcohol, drugs, self-harm, or suicidal ideation.  Focus on the reason for THIS attendance. Past history alone is insufficient unless it clearly explains the current presentation.  If a patient has left without being seen, use the triage note to determine whether the presentation qualifies.  If a patient has attended with a presentation meeting multiple criteria (e.g. under the influence of alcohol AND having taken an overdose of benzodiazepines with self-harm intent), all applicable sections should be completed. | Yes / No |
| **Alcohol related** |  | Alcohol has had a direct or contributory effect leading to the presentation (e.g. current intoxication, consequences of intoxication, organ damage). | Yes / No |
|  | Alcohol intoxication | Documentation in the notes of intoxication during the patient's time in the ED. | Yes / No |
|  | Alcohol dependence | Documentation in the notes of features of alcohol withdrawal or a requirement for monitoring/treatment for alcohol withdrawal. | Yes / No |
|  | Alcohol-related accidental injury | Documentation in the notes of an injury occurring as a consequence of alcohol intoxication, or while under the influence of alcohol. | Yes / No |
| **Drug related** |  | Drugs have had a direct or contributory effect leading to the presentation. Covers street drugs (e.g. heroin, street benzodiazepines) and psychoactive substances including prescription medications (regardless of source) which have not been used in their intended manner (e.g. codeine would be included; paracetamol used therapeutically would not). Includes harms as a consequence of drug use.  Note: If a substance is not a pharmaceutical prescribed directly to the patient, do not assume the named substance is correct (e.g. 'street valium' is rarely genuine diazepam - use 'street' prefix for any substance obtained illegally). | Yes / No |
|  | Acute drug intoxication | Documentation in the notes of drug-related intoxication during the patient's time in the ED. | Yes / No |
|  | Drug dependence | Documentation in the notes of features of drug withdrawal or a requirement for monitoring/treatment for drug withdrawal. | Yes / No |
|  | Drug-related accidental injury | Documentation in the notes of an injury occurring as a consequence of drug-related intoxication, or while under the influence of drugs. | Yes / No |
| **Self-harm / suicidal ideation related** |  | Self-harm includes both physical methods of harm (e.g. hanging, cutting) and ingestion methods (e.g. paracetamol overdose). Also includes suicidal ideation (current thoughts of wanting to die or kill oneself, even if no act has occurred). | Yes / No |
|  | Intent | Choose the single best category on the balance of probability:  1 = Suicidal ideation - thinking about killing oneself without acting on the thoughts  2 = Self-injury - non-fatal intentional injury without suicidal intent (or with unknown intent)  3 = Suicide attempt - non-fatal intentional injury with suicidal intent, regardless of likely lethality  4 = Suicide - fatal intentional injury with suicidal intent  5 = Unknown - intent cannot be determined from the note | 1–5 |
|  | Type of act | Choose the single best category on the balance of probability:  1 = Physical means only (e.g. hanging, cutting)  2 = Drug overdose with intention of self-harm  3 = Physical means and drug overdose  4 = Not disclosed | 1–4 |
| **Homelessness** |  | Is the patient homeless? Choose the single most applicable category:  1 = Living rough  2 = Living in emergency accommodation (e.g. overnight shelter)  3 = Living in temporary accommodation for the homeless (e.g. homeless hostel)  4 = Living in non-conventional dwelling due to lack of housing (e.g. mobile home, temporary structure)  5 = Doubling-up with family or friends (e.g. sofa-surfing)  6 = Other homeless situation  7 = Not homeless | 1–7 |

*Criteria: bold text denotes a top-level domain. Sub-criteria are assessed only when the parent domain criterion is marked 'Yes'. For self-harm intent and type of act, the coder selects the single best option on the balance of probability. Homelessness was extracted as part of the social history module and analysed as a binary variable (categories 1–6 = homeless; 7 = not homeless) for all performance metrics reported in the paper.*

### **Supplementary Appendix 2**

**Prompt Contract and JSON Output Schema for LLM Extraction**

**1. LLM Configuration**

Model: Llama 3.1 70B Instruct (Meta), deployed locally using vLLM (version 0.6.0) on two NVIDIA A100 GPUs within the NHS DataLoch secure data environment (Royal Infirmary of Edinburgh). Inference used deterministic decoding (temperature 0), a maximum sequence length of 8,192 tokens, and a maximum generated output of 20 tokens per task.

All runs used deterministic configuration: temperature = 0, fixed random seed, sampling disabled. These settings were applied consistently across all prompts and all records to ensure reproducibility.

The model received no fine-tuning and no retrieval-augmented generation (RAG). All prompts were applied to the raw clinical text fields only.

**2. Prompt-Contracting Methodology**

Prompts were developed using a structured contracting approach. Each prompt was iteratively developed against a held-out development set to maximise specificity of output format (single-line, machine-parseable) while preserving sensitivity to clinically relevant signals.

Key design principles applied across all prompts:

• Binary or categorical output only - no free-text explanations permitted in output

• Explicit fail condition: any output deviating from the specified format is treated as a task failure

• Conservative default: uncertainty always resolved towards 0 (negative) rather than 1 (positive)

• Current attendance focus: historical information alone is insufficient to flag a positive unless it directly explains the current presentation

• Template variable: {{text}} was substituted at runtime with concatenated clinical note fields for each record

**3. Clinical Text Fields Provided**

The following text fields from the emergency department record system were concatenated and provided to the model as the {{text}} variable for each attendance:

• Triage note

• Presenting complaint

• Clinical notes (all available entries)

• Nursing notes

• Discharge summary

• Referral letters and liaison correspondence (where present in the record)

No structured coded fields (ICD-10 codes, SNOMED codes, triage category codes) were provided. The model operated solely on free-text documentation.

**4. Processing Pipeline and Prompt Execution Order**

Records were processed in two stages:

Stage 1 - Screening: The top-level screening prompt was applied to all 2,256 records. Records returning PRESENTATION MATCHES: 0 were marked negative for all domains and subdomains and were not processed further.

Stage 2 - Domain and subdomain extraction: Records returning PRESENTATION MATCHES: 1 were passed to the alcohol, drug, and self-harm domain prompts. Records positive for a domain were then passed to the corresponding subdomain prompts. The homelessness prompt was applied to all records flagged positive by Stage 1.

*Note: Processing order within Stage 2 was parallel (domain prompts were run independently for each record). Subdomain prompts were applied conditionally on parent domain positivity.*

**5. Top-Level Domain Prompts**

**Top-level screening prompt**

*Note: Applied to all 2,256 records as the initial triage step. Records flagged as '1' were passed to the domain-specific prompts below. Records flagged as '0' were assigned negative for all domains.*

System prompt name: "Presentation of interest (if No move to next)_norm"

Your task:

Read the following clinical note and decide if the patient's attendance in the emergency

department is related to any of the following:

- ALCOHOL (current effects or consequences of alcohol use)

- DRUGS (current effects or consequences of prescription or illegal drugs used other than as

intended; exclude simple therapeutic use of over-the-counter medicines)

- SELF_HARM (any act of intentional self-harm, regardless of method)

- SUICIDAL_IDEATION (current thoughts of wanting to die or kill oneself, even if no act has

occurred)

IMPORTANT RULES:

- Focus on the reason for THIS attendance / presentation and its current effects or

consequences.

- Do not treat past history alone as sufficient unless it clearly explains the CURRENT

presentation.

- If a patient has left without being seen, use the TRIAGE NOTE or any initial documentation

to make your decision.

- Only mark 1 when the record clearly supports that the CURRENT presentation is related to

alcohol, drugs or self-harm.

- If you are unsure, use 0.

OUTPUT FORMAT (MUST BE EXACTLY ONE LINE):

PRESENTATION MATCHES: 1 or 0

CLINICAL RECORD:

{{text}}

Do not output anything else. Do not output backticks. Do not output code blocks.

Do not output explanations. Do not output extra lines.

If you output anything else, you FAIL the task.

**Alcohol domain prompt**

System prompt name: "Alcohol related?_norm"

Your task:

Read the following clinical note and determine if alcohol had a direct or contributory effect

to the admission.

Rules:

- Use 1 = yes, 0 = no.

- Only mark 1 when the record clearly supports it.

- If you are unsure, use 0.

Output MUST be EXACTLY one line ONLY in this format:

ALCOHOL: 1 or 0

CLINICAL RECORD:

{{text}}

Do not output anything else. Do not output backticks. Do not output code blocks.

Do not output explanations. Do not output extra lines.

If you output anything else, you FAIL the task.

**Drug domain prompt (primary)**

*Note: Primary drug prompt used for all reported analyses. See refined drug prompt below.*

System prompt name: "Drug related?_norm"

Your task:

Read the following clinical note and determine if drugs had a direct or contributory effect

to the admission, where the patient is taking prescription or illegal drugs (not

over-the-counter like paracetamol or ibuprofen) not for their intended purpose.

Rules:

- Use 1 = yes, 0 = no.

- Only mark 1 when the record clearly supports it.

- If you are unsure, use 0.

Output MUST be EXACTLY one line ONLY in this format:

DRUGS: 1 or 0

CLINICAL RECORD:

{{text}}

Do not output anything else. Do not output backticks. Do not output code blocks.

Do not output explanations. Do not output extra lines.

If you output anything else, you FAIL the task.

**Self-harm / suicidal ideation domain prompt**

System prompt name: "Self-harm OR suicidal ideation related?_norm"

Your task:

Read the following clinical note and determine if the admission is due to self-harm, which

includes both physical methods of harm (e.g. hanging, cutting) and ingestion methods

(e.g. paracetamol overdose).

Rules:

- Use 1 = yes, 0 = no.

- Only mark 1 when the record clearly supports it.

- If you are unsure, use 0.

Output MUST be EXACTLY one line ONLY in this format:

SELF_HARM: 1 or 0

CLINICAL RECORD:

{{text}}

Do not output anything else. Do not output backticks. Do not output code blocks.

Do not output explanations. Do not output extra lines.

If you output anything else, you FAIL the task.

**6. Subdomain Prompts**

*Subdomain prompts were applied only to records positive for the parent domain (or positive at Stage 1 screening for the homelessness prompt). Performance metrics are reported within the 287 clinician-reviewed cases.*

**Alcohol subdomain: Intoxication**

System prompt name: "Alcohol Intoxication_norm"

Your task:

Read the following clinical note and follow the instructions to decide if there is evidence

that the patient was intoxicated with alcohol during this emergency department attendance.

Examples of relevant evidence include:

- documentation of alcohol intoxication

- documentation of being drunk, intoxicated, under the influence of alcohol, or smelling

of alcohol

Rules:

- Use 1 = yes, 0 = no.

- Only mark 1 when the record clearly supports alcohol intoxication during this attendance.

- If you are unsure, use 0.

Output MUST be EXACTLY one line ONLY in this format:

ALCOHOL_INTOXICATION: 1 or 0

CLINICAL RECORD:

{{text}}

Do not output anything else. Do not output backticks. Do not output code blocks.

Do not output explanations. Do not output extra lines.

If you output anything else, you FAIL the task.

**Alcohol subdomain: Dependence**

System prompt name: "Alcohol dependence_norm"

Your task:

Read the following clinical note and follow the instructions to decide if there is evidence

of alcohol dependence or chronic harmful alcohol use.

Examples of relevant evidence include:

- a diagnosis of alcohol dependence or alcohol use disorder

- repeated heavy drinking, daily drinking, or chronic harmful use clearly described

- long-term problematic alcohol use contributing to the patient's condition

Rules:

- Use 1 = yes, 0 = no.

- Only mark 1 when the record clearly supports alcohol dependence or chronic harmful alcohol

use.

- If you are unsure, use 0.

Output MUST be EXACTLY one line ONLY in this format:

ALCOHOL_DEPENDENCE: 1 or 0

CLINICAL RECORD:

{{text}}

Do not output anything else. Do not output backticks. Do not output code blocks.

Do not output explanations. Do not output extra lines.

If you output anything else, you FAIL the task.

**Alcohol subdomain: Accidental injury**

System prompt name: "Alcohol related accidental injury_norm"

Your task:

Read the following clinical note and follow the instructions to decide if there is evidence

that the patient has an ACCIDENTAL INJURY which occurred as a consequence of alcohol

intoxication or while under the influence of alcohol.

Rules:

- Use 1 = yes, 0 = no.

- Only mark 1 when the injury is clearly linked to alcohol intoxication.

- If you are unsure, use 0.

Output MUST be EXACTLY one line ONLY in this format:

ALCOHOL_ACCIDENTAL_INJURY: 1 or 0

CLINICAL RECORD:

{{text}}

Do not output anything else. Do not output backticks. Do not output code blocks.

Do not output explanations. Do not output extra lines.

If you output anything else, you FAIL the task.

**Drug subdomain: Acute intoxication**

System prompt name: "Acute Drug Intoxication_norm"

Your task:

Read the following clinical note and follow the instructions to decide if there is evidence

the patient was acutely intoxicated with drugs during this emergency department attendance.

Rules:

- Use 1 = yes, 0 = no.

- Only mark 1 when the record clearly supports acute drug intoxication.

- If you are unsure, use 0.

Output MUST be EXACTLY one line ONLY in this format:

ACUTE_DRUG_INTOXICATION: 1 or 0

CLINICAL RECORD:

{{text}}

Do not output anything else. Do not output backticks. Do not output code blocks.

Do not output explanations. Do not output extra lines.

If you output anything else, you FAIL the task.

**Drug subdomain: Dependence**

System prompt name: "Drug dependence_norm"

Your task:

Read the following clinical note and follow the instructions to decide if there is

documentation of drug dependence.

Rules:

- Use 1 = yes, 0 = no.

- Only mark 1 when the record clearly supports drug dependence.

- If you are unsure, use 0.

Output MUST be EXACTLY one line ONLY in this format:

DRUG_DEPENDENCE: 1 or 0

CLINICAL RECORD:

{{text}}

Do not output anything else. Do not output backticks. Do not output code blocks.

Do not output explanations. Do not output extra lines.

If you output anything else, you FAIL the task.

**Drug subdomain: Accidental injury**

System prompt name: "Drug related accidental injury_norm"

Your task:

Read the following clinical note and follow the instructions to decide if there is

documentation of an accidental injury occurring as a consequence of drug use, or while

the patient was under the influence of drugs.

Rules:

- Use 1 = yes, 0 = no.

- Only mark 1 when the record clearly supports a drug-related accidental injury.

- If you are unsure, use 0.

Output MUST be EXACTLY one line ONLY in this format:

DRUG_RELATED_INJURY: 1 or 0

CLINICAL RECORD:

{{text}}

Do not output anything else. Do not output backticks. Do not output code blocks.

Do not output explanations. Do not output extra lines.

If you output anything else, you FAIL the task.

**Self-harm subdomain: Intent classification**

*Note: Applied only to records flagged as self-harm positive by the top-level self-harm prompt.*

System prompt name: "Intent_norm"

Your task:

Read the following clinical note and follow the instructions to determine the patient's

self-harm intent category.

Use exactly one of the following codes:

1 = suicidal ideation (thinking about killing oneself without acting on the thoughts)

2 = self-injury (non-fatal intentional injury without suicidal intent or with unknown intent)

3 = suicide attempt (non-fatal intentional injury with suicidal intent)

4 = suicide (fatal intentional injury with suicidal intent)

5 = unknown (intent cannot be determined from the note)

Rules:

- Output ONLY the single number (1, 2, 3, 4, or 5).

- Do NOT output any words, labels, punctuation, or explanations.

- If the intent cannot be confidently identified, choose 5.

Output MUST be EXACTLY:

<NUMBER>

CLINICAL RECORD:

{{text}}

Do not output anything else. Do not output backticks. Do not output code blocks.

Do not output explanations. Do not output extra lines.

If you output anything else, you FAIL the task.

**Self-harm subdomain: Type of act**

System prompt name: "Type of act_norm"

Your task:

Read the following clinical note and follow the instructions to determine the type of

self-harm act.

Use exactly one of the following codes:

1 = includes physical means (e.g. hanging, cutting)

2 = includes drug overdose with intention of self-harm

3 = physical means and drug overdose

4 = not disclosed

Rules:

- Output ONLY the single number: 1, 2, 3, or 4.

- Do NOT output additional text or explanations.

- If the act type is unclear or not documented, output 4.

Output MUST be EXACTLY:

<NUMBER>

CLINICAL RECORD:

{{text}}

Do not output anything else. Do not output backticks. Do not output code blocks.

Do not output explanations. Do not output extra lines.

If you output anything else, you FAIL the task.

**Homelessness (social history module)**

*Note: For performance analysis, homelessness was analysed as a binary variable. A categorical prompt (codes 1–7) was also run to characterise type of homelessness; this is not reported in the current manuscript.*

System prompt name: "Homeless_YESNO"

Your task:

Read the following clinical note and determine if there is evidence of the patient being

homeless.

Rules:

- Use 1 = yes, 0 = no.

- Only mark 1 when clearly documented.

- If unsure, use 0.

Output MUST be EXACTLY:

HOMELESS_YESNO: 1 or 0

CLINICAL RECORD:

{{text}}

Do not output anything else. Do not output code blocks. Do not output backticks.

Do not output explanations. Do not output extra lines.

If you output anything else, you FAIL the task.

**7. Refined Drug Domain Prompt (Post Hoc, Not Used in Reported Analyses)**

Following analysis of the primary drug prompt's high false-positive rate (PPV 0.370), a refined prompt was developed post hoc to investigate whether precision could be improved. The refined prompt incorporated additional exclusion criteria based on the three systematic error sources identified on post hoc review:

(1) Adverse medication reactions documented in the notes

(2) Composite triage codes that included drug-related terms without genuine drug involvement

(3) Therapeutic benzodiazepine administration for alcohol withdrawal management

The refined prompt worsened balanced accuracy from 0.959 to 0.755 due to 41 additional false negatives; the primary prompt is therefore used for all reported results. Rogan–Gladen adjustment of the primary prompt is preferred for prevalence estimation.

System prompt name: "Drug related? (refined, post hoc)_norm"

Your task:

Read the following clinical note and determine if drugs had a direct or contributory effect

to the admission, where the patient is taking prescription or illegal drugs (not

over-the-counter like paracetamol or ibuprofen) not for their intended purpose.

Do NOT mark 1 (treat as 0) when the ONLY drug involvement is one of the following:

- an adverse reaction to a medication taken as prescribed or as directed

- a triage or presenting-complaint code that mentions drugs, but where the clinical

notes do not support genuine drug involvement in this attendance

- benzodiazepines or other medication given by staff to treat alcohol withdrawal

Rules:

- Use 1 = yes, 0 = no.

- Only mark 1 when the record clearly supports genuine, non-therapeutic drug involvement.

- If you are unsure, use 0.

Output MUST be EXACTLY one line ONLY in this format:

DRUGS: 1 or 0

CLINICAL RECORD:

{{text}}

Do not output anything else. Do not output backticks. Do not output code blocks.

Do not output explanations. Do not output extra lines.

If you output anything else, you FAIL the task.

**8. Output Parsing and Quality Control**

All prompt outputs were parsed programmatically.

### **Supplementary Appendix 3**

**Full Confusion Matrices - All Domains and Subdomains**

This appendix presents (a) top-level 2×2 confusion matrices for each domain × strategy combination (N=2,256); (b) multi-class confusion matrices for self-harm intent and type of act for clinician review and LLM vs gold standard (n=87); and (c) subdomain performance metrics for all binary subdomains assessed within the 287 clinician-reviewed cases.

*Row 0 in multi-class matrices denotes cases in which the rater did not identify the presentation as within scope for the self-harm domain.*

**(a) Top-Level 2×2 Confusion Matrices (N=2,256)**

*TP = true positive; FP = false positive; FN = false negative; TN = true negative. Green cell = concordant with gold standard; red cell = discordant.*

**Alcohol Domain**

***Clinical Coding***

|  |  | **Gold Standard** | |  |
| --- | --- | --- | --- | --- |
|  |  | **Positive** | **Negative** | **Total** |
| **Clinical Coding** | **Positive** | 46 | 6 | 52 |
|  | **Negative** | 129 | 2075 | 2204 |
| **Total** |  | **175** | **2081** | **2256** |

***Clinician Review***

|  |  | **Gold Standard** | |  |
| --- | --- | --- | --- | --- |
|  |  | **Positive** | **Negative** | **Total** |
| **Clinician Review** | **Positive** | 151 | 8 | 159 |
|  | **Negative** | 24 | 2073 | 2097 |
| **Total** |  | **175** | **2081** | **2256** |

***LLM (Llama 3.1 70B Instruct)***

|  |  | **Gold Standard** | |  |
| --- | --- | --- | --- | --- |
|  |  | **Positive** | **Negative** | **Total** |
| **LLM** | **Positive** | 155 | 2 | 157 |
|  | **Negative** | 20 | 2079 | 2099 |
| **Total** |  | **175** | **2081** | **2256** |

**Drug Domain**

***Clinical Coding***

|  |  | **Gold Standard** | |  |
| --- | --- | --- | --- | --- |
|  |  | **Positive** | **Negative** | **Total** |
| **Clinical Coding** | **Positive** | 30 | 27 | 57 |
|  | **Negative** | 61 | 2138 | 2199 |
| **Total** |  | **91** | **2165** | **2256** |

***Clinician Review***

|  |  | **Gold Standard** | |  |
| --- | --- | --- | --- | --- |
|  |  | **Positive** | **Negative** | **Total** |
| **Clinician Review** | **Positive** | 53 | 1 | 54 |
|  | **Negative** | 38 | 2164 | 2202 |
| **Total** |  | **91** | **2165** | **2256** |

***LLM (Llama 3.1 70B Instruct)***

|  |  | **Gold Standard** | |  |
| --- | --- | --- | --- | --- |
|  |  | **Positive** | **Negative** | **Total** |
| **LLM** | **Positive** | 90 | 153 | 243 |
|  | **Negative** | 1 | 2012 | 2013 |
| **Total** |  | **91** | **2165** | **2256** |

*Note: the LLM's near-perfect sensitivity (0.989) but low PPV (0.370) for the drug domain reflects three systematic false positive sources identified on post hoc review: adverse medication reactions, composite triage codes, and therapeutic benzodiazepine administration for alcohol withdrawal. Rogan–Gladen adjustment of apparent prevalence (10.8%) yields an adjusted estimate of 4.1% [95% CI 2.9–5.2%], consistent with the gold-standard prevalence of 4.0%.*

**Self-Harm Domain**

***Clinical Coding***

|  |  | **Gold Standard** | |  |
| --- | --- | --- | --- | --- |
|  |  | **Positive** | **Negative** | **Total** |
| **Clinical Coding** | **Positive** | 33 | 4 | 37 |
|  | **Negative** | 62 | 2157 | 2219 |
| **Total** |  | **95** | **2161** | **2256** |

***Clinician Review***

|  |  | **Gold Standard** | |  |
| --- | --- | --- | --- | --- |
|  |  | **Positive** | **Negative** | **Total** |
| **Clinician Review** | **Positive** | 78 | 9 | 87 |
|  | **Negative** | 17 | 2152 | 2169 |
| **Total** |  | **95** | **2161** | **2256** |

***LLM (Llama 3.1 70B Instruct)***

|  |  | **Gold Standard** | |  |
| --- | --- | --- | --- | --- |
|  |  | **Positive** | **Negative** | **Total** |
| **LLM** | **Positive** | 92 | 11 | 103 |
|  | **Negative** | 3 | 2150 | 2153 |
| **Total** |  | **95** | **2161** | **2256** |

**(b) Multi-Class Confusion Matrices - Self-Harm Intent and Type of Act**

To prevent identification of individuals within these small self-harm subgroups, all cell and marginal counts in the four matrices below were subject to statistical disclosure control, and are shown as an upper bound rather than an exact figure. Each non-zero count is given as the next multiple of five above the true value (so <5 denotes a count of 1 to 4, <10 denotes 5 to 9, <15 denotes 10 to 14, and so on). Suppression is applied to every non-zero figure, including the larger cells and the row and column subtotals, and not only to the smallest cells. This is deliberate, because if the larger cells or the subtotals were shown exactly, the suppressed small cells could be recovered by back-calculation. True zero counts are shown as 0, and the overall total of 87 is retained, as neither discloses any individual-level information.

Assessed within n=87 gold-standard self-harm positive cases. Note: of the 95 gold-standard self-harm cases, 87 were identified by the clinician reviewer and therefore have subdomain classification data; the remaining 8 were identified by the LLM only and lack subdomain data. Category 0 in these matrices denotes cases where the rater did not identify the presentation as self-harm at the top-level domain - i.e. a false negative at the domain level, coded as 0 to allow kappa to penalise missed cases. The LLM has no row-0 entries (it identified all 87 at the top level); the clinician has several (cases missed at the domain level).

*Policy implication: of the 87 classified cases, 16 (18.4%) were potentially suitable for mental health crisis centre streaming (13 suicidal ideation; 3 unknown intent without overdose or physical act). The 8 LLM-only cases could not be classified for streaming eligibility.*

*Rows = rater's classification; columns = gold-standard classification. Diagonal cells represent agreement; off-diagonal cells represent errors.*

**Self-Harm Intent - Clinician Review vs Gold Standard**

Intent categories: 0 = not identified as self-harm; 1 = suicidal ideation; 2 = self-injury; 3 = suicide attempt; 4 = completed suicide; 5 = unknown intent

*Observed agreement: 0.575. Expected agreement: 0.238. Cohen's κ = 0.442 [95% CI 0.325, 0.559]. Weighted κ = 0.309 [0.155, 0.469].*

|  | **Gold Standard** | | | | | | |
| --- | --- | --- | --- | --- | --- | --- | --- |
| **Clinician** | **0 (not flagged)** | **1 (suicidal ideation)** | **2 (self-injury)** | **3 (suicide attempt)** | **4 (completed suicide)** | **5 (unknown)** | **Total** |
| **0 (not flagged)** | 0 | <5 | <5 | <5 | 0 | <10 | **<10** |
| **1 (suicidal ideation)** | 0 | <15 | <5 | <10 | 0 | 0 | **<20** |
| **2 (self-injury)** | 0 | 0 | <10 | <5 | 0 | 0 | **<10** |
| **3 (suicide attempt)** | 0 | 0 | 0 | <30 | 0 | 0 | **<30** |
| **4 (completed suicide)** | 0 | 0 | 0 | <5 | 0 | 0 | **<5** |
| **5 (unknown)** | 0 | 0 | <15 | <5 | <5 | <5 | **<25** |
| **Total** | **0** | **<15** | **<25** | **<45** | **<5** | **<10** | **87** |

**Self-Harm Intent - LLM vs Gold Standard**

Intent categories: 0 = not identified as self-harm; 1 = suicidal ideation; 2 = self-injury; 3 = suicide attempt; 4 = completed suicide; 5 = unknown intent

*Observed agreement: 0.793. Expected agreement: 0.399. Cohen's κ = 0.656 [95% CI 0.514, 0.787]. Weighted κ = 0.595 [0.423, 0.750].*

|  | **Gold Standard** | | | | | | |
| --- | --- | --- | --- | --- | --- | --- | --- |
| **LLM** | **0 (not flagged)** | **1 (suicidal ideation)** | **2 (self-injury)** | **3 (suicide attempt)** | **4 (completed suicide)** | **5 (unknown)** | **Total** |
| **0 (not flagged)** | 0 | 0 | 0 | 0 | 0 | 0 | **0** |
| **1 (suicidal ideation)** | 0 | <10 | 0 | 0 | 0 | 0 | **<10** |
| **2 (self-injury)** | 0 | 0 | <20 | 0 | 0 | <5 | **<20** |
| **3 (suicide attempt)** | 0 | <10 | <10 | <45 | 0 | <5 | **<60** |
| **4 (completed suicide)** | 0 | 0 | 0 | 0 | <5 | 0 | **<5** |
| **5 (unknown)** | 0 | 0 | 0 | 0 | 0 | <5 | **<5** |
| **Total** | **0** | **<15** | **<25** | **<45** | **<5** | **<10** | **87** |

**Self-Harm Type of Act - Clinician Review vs Gold Standard**

Type categories: 0 = not identified as self-harm; 1 = physical means (e.g. hanging, cutting); 2 = drug overdose with intention of self-harm; 3 = physical means and drug overdose; 4 = not disclosed

*Observed agreement: 0.816. Expected agreement: 0.406. Cohen's κ = 0.690 [95% CI 0.565, 0.809]. Weighted κ = 0.600 [0.418, 0.770].*

|  | **Gold Standard** | | | | | |
| --- | --- | --- | --- | --- | --- | --- |
| **Clinician** | **0 (not flagged)** | **1 (physical means)** | **2 (drug overdose)** | **3 (physical + overdose)** | **4 (not disclosed)** | **Total** |
| **0 (not flagged)** | 0 | <5 | <5 | 0 | <5 | **<15** |
| **1 (physical means)** | 0 | <15 | 0 | 0 | 0 | **<15** |
| **2 (drug overdose)** | 0 | 0 | <50 | <5 | 0 | **<55** |
| **3 (physical + overdose)** | 0 | 0 | 0 | <5 | 0 | **<5** |
| **4 (not disclosed)** | 0 | <5 | 0 | 0 | <10 | **<10** |
| **Total** | **0** | **<20** | **<55** | **<10** | **<15** | **87** |

**Self-Harm Type of Act - LLM vs Gold Standard**

Type categories: 0 = not identified as self-harm; 1 = physical means; 2 = drug overdose with intention of self-harm; 3 = physical means and drug overdose; 4 = not disclosed

*Observed agreement: 0.885. Expected agreement: 0.466. Cohen's κ = 0.785 [95% CI 0.652, 0.898]. Weighted κ = 0.699 [0.524, 0.860].*

|  | **Gold Standard** | | | | | |
| --- | --- | --- | --- | --- | --- | --- |
| **LLM** | **0 (not flagged)** | **1 (physical means)** | **2 (drug overdose)** | **3 (physical + overdose)** | **4 (not disclosed)** | **Total** |
| **0 (not flagged)** | 0 | 0 | 0 | 0 | 0 | **0** |
| **1 (physical means)** | 0 | <20 | 0 | 0 | <5 | **<20** |
| **2 (drug overdose)** | 0 | <5 | <55 | 0 | <10 | **<65** |
| **3 (physical + overdose)** | 0 | 0 | 0 | <10 | 0 | **<10** |
| **4 (not disclosed)** | 0 | 0 | 0 | 0 | <5 | **<5** |
| **Total** | **0** | **<20** | **<55** | **<10** | **<15** | **87** |

**(c) Subdomain Performance Metrics (n=287)**

Assessed within the 287 cases flagged for review by any strategy. Performance metrics are conditional on parent domain positivity (i.e. the denominator for each subdomain is the number of cases within the 287 that were positive for the parent domain by the gold standard). For homelessness, the denominator is all 287 cases flagged at Stage 1.

*BA = balanced accuracy = (sensitivity + specificity) / 2. F1 = harmonic mean of sensitivity and PPV.*

| **Subdomain** | **Clinician** | | | | **LLM** | | | |
| --- | --- | --- | --- | --- | --- | --- | --- | --- |
|  | **Sens** | **Spec** | **BA** | **F1** | **Sens** | **Spec** | **BA** | **F1** |
| Alcohol: Intoxication | 0.955 | 0.727 | 0.841 | 0.926 | 0.928 | 0.864 | 0.896 | 0.936 |
| Alcohol: Dependence | 0.892 | 0.978 | 0.935 | 0.928 | 0.877 | 0.967 | 0.922 | 0.912 |
| Alcohol: Injury | 0.759 | 0.928 | 0.843 | 0.807 | 0.879 | 0.969 | 0.924 | 0.911 |
| Drug: Intoxication | 0.053 | 0.727 | 0.390 | 0.087 | 0.947 | 0.273 | 0.610 | 0.800 |
| Drug: Dependence | 0.000 | 0.882 | 0.441 | 0.000 | 1.000 | 0.118 | 0.559 | 0.571 |
| Drug: Injury | 0.200 | 0.615 | 0.408 | 0.235 | 0.800 | 0.385 | 0.592 | 0.615 |
| Homelessness | 0.733 | 0.982 | 0.857 | 0.733 | 0.800 | 0.982 | 0.891 | 0.774 |

*Bootstrap 95% CIs (10,000 resamples) for balanced accuracy are reported in Supplementary Table S8. Sens = sensitivity; Spec = specificity; BA = balanced accuracy; F1 = F1 score.*

*Drug subdomain note: clinician sensitivity for acute drug intoxication (BA 0.390) and drug dependence (BA 0.441) was low, while LLM sensitivity was high but specificity poor. This pattern reflects a documentation artefact: intoxication and dependence in drug presentations are frequently implied rather than explicitly stated in ED notes. Clinicians tended to record the documented presentation rather than infer subdomain classifications, depressing sensitivity. The LLM applied definitions more literally to contextual cues, recovering true positives at the cost of false positives. These metrics should be interpreted cautiously as reflecting documentation norms rather than the intrinsic capability of either approach.*
