## Supplementary material for "Half of alcohol, drug, and self-harm presentations cannot be identified in coded emergency department data: a diagnostic accuracy study of a large language model": STARD-AI

| **Item** | **Recommendation** | **Location in manuscript** | **Reported / N/A** |
| --- | --- | --- | --- |
| **Title or abstract** | | | |
| 1 | Identification as a study of AI-centred diagnostic accuracy, reporting at least one measure of accuracy in the title or abstract | Title; Abstract (p1-2) | Reported |
| **Abstract** | | | |
| 2 | Structured summary of study design, methods, results and conclusions | Abstract (p2) | Reported |
| **Introduction** | | | |
| 3 | Scientific and clinical background, intended use of the index test, whether novel or established, and its integration into a workflow | Introduction (p3) | Reported |
| 4 | Study objectives and hypotheses | Introduction (p3) | Reported |
| **Methods - Study design** | | | |
| 5 | Whether data collection was planned before (prospective) or after (retrospective) the index test and reference standard | Methods: Study design and setting (retrospective); Supplementary Figure S1 (two-phase study design) (p4) | Reported |
| **Methods - Ethics** | | | |
| 6 | Formal approval from an ethics committee; if not required, justify | Methods: Ethical approval (p4) | Reported |
| **Methods - Participants** | | | |
| 7 | Eligibility criteria: inclusion and exclusion criteria at participant and data level | Methods: Study design and setting; Appendix 1 (p4) | Reported |
| 8 | On what basis potentially eligible participants were identified | Methods: Study design and setting (p4) | Reported |
| 9 | Where and when potentially eligible participants were identified (setting, location, dates) | Methods: Study design and setting (p4) | Reported |
| 10 | Whether participants formed a consecutive, random, or convenience series | Methods: Study design and setting (consecutive attendances) (p4) | Reported |
| **Methods - Dataset** | | | |
| 11 | Source of the data and whether routinely collected, collected for the study, or from an open-source repository | Methods: Study design and setting (routine ED records via DataLoch) (p4) | Reported |
| 12 | Who undertook the annotations (experience, background) and how | Methods: Reference standard; Appendix 1 (p5) | Reported |
| 13 | Devices/software (with version) used to engineer the index test, and its intended use | Methods: Classification strategies; Appendix 2 (Llama 3.1 70B Instruct; vLLM 0.6.0; DataLoch TRE) (p4) | Reported |
| 14 | Data acquisition protocols and pre-processing in sufficient detail to allow replication | Methods: Classification strategies; Appendix 2 (p4) | Reported |
| **Methods - Test methods** | | | |
| 15a | Index test, in sufficient detail to allow replication | Methods: Classification strategies; Appendix 2 (p4) | Reported |
| 15b | How the index test was developed (training, validation, testing, external evaluation; sample sizes) | Methods: Classification strategies; Appendix 2 (prompt contracting on a held-out development set; no fine-tuning) | Reported |
| 15c | Definition of and rationale for test positivity cut-offs or result categories, pre-specified vs exploratory | Methods: Classification strategies; Outcomes (pre-specified binary outputs) | Reported |
| 15d | The specified end user of the index test and the expertise required | Methods; Discussion: existing governance (automated extraction within the TRE) (p4, p12) | Reported |
| 16a | Reference standard, in sufficient detail to allow replication | Methods: Reference standard (p5) | Reported |
| 16b | Rationale for choosing the reference standard | Methods: Reference standard (p5) | Reported |
| 16c | Definition of and rationale for reference-standard positivity cut-offs or categories | Methods: Reference standard; Outcomes; Appendix 1 (p5) | Reported |
| 17a | Whether clinical information and reference-standard results were available to index-test performers | Methods: Classification strategies (LLM used free text only; blind to reference standard) (p4) | Reported |
| 17b | Whether clinical information and index-test results were available to reference-standard assessors | Methods: Reference standard (clinicians blind to coding and LLM output) (p5) | Reported |
| **Methods - Analysis** | | | |
| 18 | Methods for estimating or comparing measures of diagnostic accuracy | Methods: Statistical analysis (p6) | Reported |
| 19 | How indeterminate index-test or reference-standard results were handled | Methods: Statistical analysis; Appendix 2 (format-failure and conservative-default rules) (p6) | Reported |
| 20 | How missing data were handled | Methods: Statistical analysis (complete text fields; no missing data) (p6) | Reported |
| 21 | Any analyses of variability in accuracy, pre-specified vs exploratory | Methods: Statistical analysis (subdomain and co-occurrence analyses pre-specified as exploratory) (p6) | Reported |
| 22 | Intended sample size and how it was determined | Methods: Study design and setting (one consecutive week; pragmatic census) (p4) | Reported |
| 23 | Details of any performance error analysis and algorithmic bias/fairness assessment | Results: Characterisation and correction of LLM error; Discussion: Limitations (formal fairness assessment not possible: no demographic linkage under the approval) (p8, p12) | Reported |
| **Results - Participants and dataset** | | | |
| 24 | Flow of participants, using a diagram | Methods; Results; Supplementary Figure S1 (two-phase study design). Participant numbers reported in text (2,256 validation week; 105,096 deployment year); no separate participant-flow diagram | Reported |
| 25 | Baseline demographic, clinical and technical characteristics | Table 1 (clinical case-mix; demographic attributes not linked under the ethics approval) (p7) | Reported |
| 26a | Distribution of severity of disease in those with the target condition | Results: Subdomain classification; Table S8 (p9) | Reported |
| 26b | Distribution of alternative diagnoses in those without the target condition | N/A - binary domain involvement; no differential-diagnosis structure | N/A |
| 27 | Time interval and clinical interventions between index test and reference standard | N/A - index test and reference standard derived from the same attendance record; no interval | N/A |
| 28 | Whether datasets represent the target-condition distribution expected in the intended-use population | Discussion: generalisability; deployment year (105,096 attendances) (p10, p12) | Reported |
| 29 | For external evaluation, how the dataset differs from training/validation/test sets | N/A - single-site study; no external evaluation dataset | N/A |
| **Results - Test results** | | | |
| 30 | Cross-tabulation of index-test results by reference-standard results | Supplementary Table S3; Appendix 3 | Reported |
| 31 | Estimates of diagnostic accuracy and their precision (e.g. 95% CI) | Table 2; Table 3; Supplementary Table S3 (p8, p11) | Reported |
| 32 | Any adverse events from the index test or reference standard | N/A - non-interventional, records-based study; no adverse events | N/A |
| **Discussion** | | | |
| 33 | Study limitations, including sources of bias, statistical uncertainty and generalisability | Discussion: Limitations (p12) | Reported |
| 34 | Implications for practice, including intended use and clinical role of the index test | Discussion: Implications for mental health crisis centres; for research and surveillance (p11-12) | Reported |
| 35 | Ethical considerations, adherence to ethical standards, and issues of fairness | Discussion: unstructured text to structured data within existing governance; Limitations (p12) | Reported |
| **Other information** | | | |
| 36 | Registration number and name of registry | N/A - observational secondary-data analysis; not registered | N/A |
| 37 | Where the full study protocol can be accessed | Data availability (p14) | Reported |
| 38 | Sources of funding and other support; role of funders | Funding (p14) | Reported |
| 39 | Commercial interests, if applicable | Competing interests (p14) | Reported |
| 40a | Availability of datasets and code; restrictions on reuse | Data availability; Appendix 2 (p14) | Reported |
| 40b | Whether outputs are stored, auditable and available for evaluation | Data availability; Discussion: existing governance (DataLoch TRE audit trail) (p12, p14) | Reported |
