## Supplementary material for "Half of alcohol, drug, and self-harm presentations cannot be identified in coded emergency department data: a diagnostic accuracy study of a large language model": STROBE

| **Item** | **Recommendation** | **Location in manuscript** | **Reported / N/A** |
| --- | --- | --- | --- |
| 1a | Title and abstract — study design | Title (p1) | Reported |
| 1b | Abstract — results summary | Abstract - structured summary (p2) | Reported |
| 2 | Introduction: background and rationale | Introduction (p3) | Reported |
| 3 | Introduction: objectives | Introduction - objectives, final paragraph (p3) | Reported |
| 4 | Study design | Methods: Study design and setting; Supplementary Figure S1 (two-phase study design) (p4) | Reported |
| 5 | Setting | Methods: Study design and setting (p4) | Reported |
| 6a | Participants — eligibility criteria | Methods: Study design and setting (p4); Appendix 1 | Reported |
| 6b | Participants — sources and selection | Methods: Study design and setting (p4) | Reported |
| 7 | Variables | Methods: Outcomes (p5); Appendix 1 | Reported |
| 8 | Data sources / measurement | Methods: Classification strategies and Reference standard (p4-5); Appendix 2 | Reported |
| 9 | Bias | Methods: Reference standard (p5) | Reported |
| 10 | Study size | Methods: Study design and setting (p4) | Reported |
| 11 | Quantitative variables | Methods: Statistical analysis (p6) | Reported |
| 12a | Statistical methods — main analyses | Methods: Statistical analysis (p6) | Reported |
| 12b | Statistical methods — subgroup analyses | Methods: Statistical analysis (p6) | Reported |
| 12c | Statistical methods — missing data | N/A - complete text fields, no missing data | N/A |
| 12d | Statistical methods — sensitivity analyses | Results: Characterisation and correction of LLM error (p8); Supplementary Table S5 | Reported |
| 12e | Statistical methods — confounding | N/A - no confounder adjustment | N/A |
| 13a | Participants — numbers in each stage | Results: Validation week (p7) and Deployment year (p10) | Reported |
| 13b | Participants — flow diagram | Participant numbers given in text (Methods p4; Results p7, p10); two-phase design in Supplementary Figure S1; no separate participant-flow diagram | — |
| 14a | Descriptive data — characteristics | Table 1 (p7) | Reported |
| 14b | Descriptive data — missing data | N/A - no missing data | N/A |
| 15 | Outcome data | Tables 1-3 (p7-8, p11); Results (p7-11) | Reported |
| 16a | Main results — unadjusted estimates | Table 3 (p11); Results: Adjusted annual prevalence (p10) | Reported |
| 16b | Main results — adjusted estimates | N/A - no model-adjusted estimates | N/A |
| 16c | Main results — absolute risk where relevant | Results: Adjusted annual prevalence and prevalence gap (p10) | Reported |
| 17 | Other analyses | Results: Subdomain classification (p9); Figure 2 (p9); Supplementary Table S8 | Reported |
| 18 | Key results | Discussion: Principal findings (p11) | Reported |
| 19 | Limitations | Discussion: Limitations (p12) | Reported |
| 20 | Interpretation | Discussion (p11-12) | Reported |
| 21 | Generalisability | Discussion: Limitations - generalisability (p12) | Reported |
| 22 | Funding | Funding (p14) | Reported |
